# Dampened inflammatory signalling and myeloid-derived suppressor-like cell accumulation reduces circulating monocytic HLA-DR density and associates with malignancy risk in long-term renal transplant recipients

**DOI:** 10.1101/2022.03.23.22272699

**Authors:** Matthew J Bottomley, Paul N Harden, Kathryn J Wood, Joanna Hester, Fadi Issa

**Author notes:** **Correspondence:** Matthew Bottomley. Equal contribution and senior authorship (Joanna Hester, Fadi Issa).

## Abstract

**Background:** Malignancy is a major cause of morbidity and mortality in transplant recipients. Identification of those at highest risk could facilitate pre-emptive intervention such as reduction of immunosuppression. Reduced circulating monocytic HLA-DR density is a marker of immune depression in the general population and associates with poorer outcome in critical illness. It has recently been used as a safety marker in adoptive cell therapy trials in renal transplantation. Despite its potential as a marker of dampened immune responses, factors that impact upon monocytic HLA-DR density and the long-term clinical sequelae of this have not been assessed in transplant recipients.

**Methods:** A cohort study of stable long-term renal transplant recipients was undertaken. Serial circulating monocytic HLA-DR density and other leucocyte populations were quantified by flow cytometry. Gene expression of monocytes was performed using the Nanostring nCounter platform, and 13-plex cytokine bead array used to quantify serum concentrations. The primary outcome was malignancy development during one-year follow-up. Risk of malignancy was calculated by univariate and multivariate proportionate hazards modelling with and without adjustment for competing risks.

**Results:** Monocytic HLA-DR density was stable in long-term renal transplant recipients (n=135) and similar to non-immunosuppressed controls (n=29), though was suppressed in recipients receiving prednisolone. Decreased mHLA-DRd was associated with accumulation of CD14+CD11b+CD33+HLA-DRlo monocytic myeloid-derived suppressor-like cells. Pathway analysis revealed downregulation of pathways relating to cytokine and chemokine signalling in monocytes with low HLA-DR density; however serum concentrations of major cytokines did not differ between these groups. There was an independent increase in malignancy risk during follow-up with decreased HLA-DR density.

**Conclusions:** Dampened chemokine and cytokine signalling drives a stable reduction in monocytic HLA-DR density in long-term transplant recipients and associates with subsequent malignancy risk. This may function as a novel marker of excess immunosuppression. Further study is needed to understand the mechanism behind this association.

## 1 Introduction

Malignancy is a leading cause of morbidity and premature mortality in long-term renal transplant recipients (RTR) (1). Overall cancer incidence is three times that of the general population, though certain tumours are significantly overrepresented, such as cutaneous squamous cell carcinoma (cSCC), with up to 250-fold excess incidence (2-6). Malignancy outcomes are poorer in transplant recipients compared to the general population and increases the subsequent risk of death with a functioning graft (DWFG) ten-fold, and a 50% chance of DWFG in the two years following cancer diagnosis, which has not improved over the last thirty years (7-9).

Monocytes are circulating precursors to macrophages, myeloid-derived suppressor cells (MDSC) and a subset of dendritic cells. These bridge innate and adaptive immune responses, partly through presentation of self and non-self peptide to CD8^+^ and CD4^+^ T cells on Major Histocompatibility Complex (MHC) Classes I and II, respectively. The most clinically relevant class II complex in transplantation is Human Leucocyte Antigen – DR (HLA-DR), which is constitutively expressed on antigen presenting cells, including monocytes, and is critical for initiation of T cell alloresponses (10).

Acute pathophysiological downregulation of circulating monocytic HLA-DR density (mHLA-DRd) has been demonstrated in a number of critical illnesses, including polytrauma (11), sepsis (12), major abdominal and cardiac surgery (13-15), acute pancreatitis (16), and severe COVID-19 infection (17). Reduced mHLA-DRd is associated with impaired initiation of adaptive immune responses *in vitro* and has been associated with poorer outcomes including enhanced risk of post-operative surgical complications, secondary/nosocomial infection and increased mortality (12-15, 17). More recently, peri-operative mHLA-DRd quantification has been used as a safety measure for both subclinical immunodepression or immune activation in trials of autologous regulatory cell infusion following organ transplantation (18-20). mHLA-DRd is reduced and may predict short-term risk of infection after lung and liver transplant; this is therefore presumed to represent a dynamic marker of excess immunosuppression (21-24). Whether this marker is stable over longer periods and correlates to long-term risk of complications of excess immunosuppression is unknown.

Historically HLA-DR density on circulating monocytes was assessed as a proportion of positive cells or by using intensity of fluorescent staining – both of which lead to challenges in standardisation and inconsistency of clinical outcomes. Subsequently a more reproducible method of HLA-DR quantification using standard curve generation was described (25). The data here reports the results of a pre-specified exploratory analysis undertaken as part of a prospective cohort study assessing immune phenotype and malignancy risk in long-term RTR. We investigated the association of clinical and immunological parameters upon mHLA-DRd, with the hypothesis that it would be reduced compared to healthy controls. Furthermore, we assessed the clinical impact of this reduction, as a potential measure of ‘excess’ immunosuppression, hypothesizing this would predict increased malignancy risk.

## 2 Materials and methods

The conduct of the study was reviewed and approved by NHS Research Ethical Committee (REC) prior to commencement (reference: 12/WS/0288) and was conducted according to the principles of the Declaration of Helsinki. Participants provided written informed consent prior to enrolment.

The study is reported according to ‘strengthening the reporting of observation studies in epidemiology’ (STROBE) guidelines (26). A completed checklist is provided in Supplementary Data.

### 2.1 Participant recruitment & clinical data collection

RTR with stable graft function and without history of malignancy (other than keratinocyte cancer), human immunodeficiency virus (HIV) or active, chronic viral hepatitis within the last five years were recruited during routine transplant outpatient follow-up. Transplant recipients with at least five years cumulative immunosuppression were recruited, or a previous history of cSCC, in order to identify a high-risk population for outcome events during follow-up. As this was an exploratory analysis, no *a priori* power calculation was undertaken.

Non-immunosuppressed controls were recruited from two sources: (a) kidney transplant (living) donor follow-up clinic (n=9) and (b) dermatology clinic, from non-immunosuppressed controls with a recent diagnosis of (fully excised) cutaneous squamous cell carcinoma (n=20). Living donor controls were recruited based on their having previously donated a kidney to a transplant recipient within the study, whilst a convenience sample of eligible controls from dermatology clinic was recruited over a six month period. Potential controls were excluded if non-Caucasian, there was a history of non-keratinocyte malignancy within the last five years or were receiving systemic immunosuppression. Non-immunosuppressed SCC controls were felt to represent the closest match to long-term transplant recipients in terms of malignancy risk, without an active malignancy at time of sampling. No difference was seen between the two recruitment sources in terms of mHLA-DRd (data not shown) and so data is reported as one group.

A questionnaire was completed at time of recruitment to assess lifetime sun exposure, family history of malignancy and smoking history, and skin type was assessed. Chronic ultraviolet radiation (UVR) exposure was assessed as described previously (3).

Clinical data were collected from medical and transplant records and pathology databases. Estimated glomerular filtration rate (eGFR) was calculated using the four-variable Modified Diet in Renal Disease equation.

### 2.2 Flow cytometry

Venepuncture was undertaken at trough levels of immunosuppression. Participants were further sampled at first routine outpatient follow-up after six months and twelve months post-recruitment.

#### 2.2.1 Monocyte HLA-DR Staining & Standard Curve Generation

Staining was undertaken as per manufacturer’s recommendations (Becton Dickinson, Wokingham, UK) and as previously published (25). Briefly, whole blood was stained with antibodies to a non-polymorphic HLA-DR epitope and ‘anti-Monocyte PerCP-Cy5.5’, followed by erythrocyte lysis and peripheral blood mononuclear cell (PBMC) fixation. The latter antibody is specific for CD14 (clone MφP9) but also non-specifically binds CD64 (FcγRI) due to the affinity of PerCP-Cy5.5 for this receptor, ensuring detection of all circulating monocytes (27) (Figure 1A). The HLA-DR – PE antibody is supplied at >95% 1:1 PE:mAb ratio.

**Figure 1:**
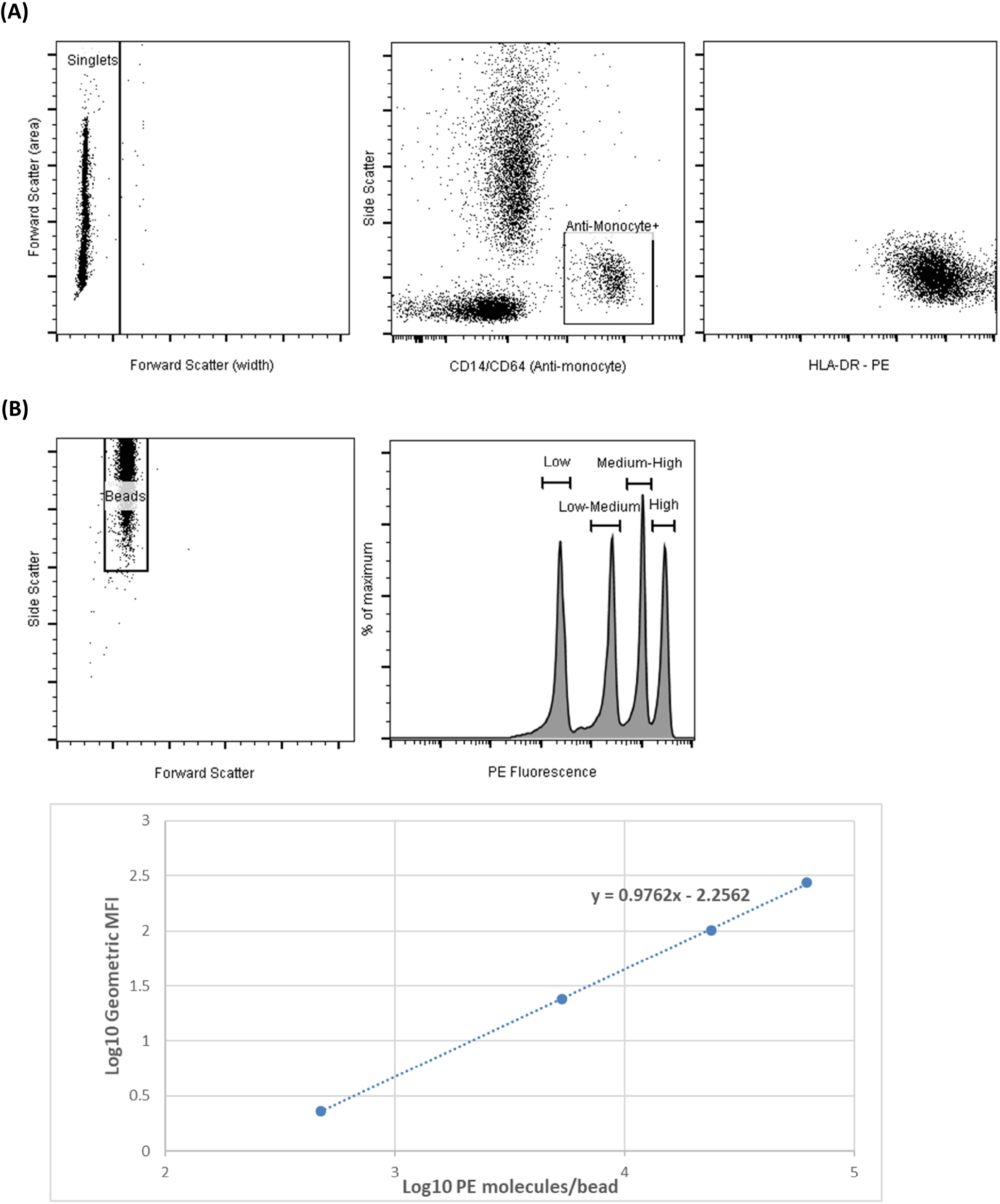
Example of HLA-DR antibody binding density calculation. (A) Illustrative staining demonstrating monocyte gating based on ‘anti-monocyte’ and HLA-DR staining of singlets; (B) example of generation of a ‘standard curve’ created by use of standardised beads with low, medium-low, medium-high and high density of phycoerythrin (PE) molecules per bead. The log-transformed number of PE molecules and log-transformed geometric mean fluorescence intensity is then plotted to generate a standard curve, from which the median number of HLA-DR antibodies (conjugated 1:1 with PE) bound per monocyte can be calculated.

All samples were analysed within four hours of venepuncture and blood was strictly stored on ice in the interim to prevent *ex vivo*, artefactual, mHLA-DRd upregulation (Supplementary Figure 1).

Reconstituted lyophilised ‘Quantibrite’ beads, containing beads conjugated with one of four pre-specified densities of PE, were used to generate standard curves of geometric mean fluorescence intensity (MFI) versus PE molecule number using linear regression (Figure 1B), yielding an equation in the format y = mx + c. The number of HLA-DR antibodies bound per cell (HLA-DR binding density, termed ‘HLA-DR density’), assuming 1:1 rato with PE molecules, was calculated using the standard curve.

#### 2.2.2 Staining of other leucocyte populations

PBMCs were isolated by density-gradient centrifugation and stained using a cocktail of antibodies (Supplementary Table 1). All flow cytometry data were acquired using a Navios flow cytometer and analyzed using Kaluza version 1.4 (both Beckman Coulter, Wycombe, UK) and FlowJoX (TreeStar, Inc) as previously published (28). Monocyte populations were stained using thawed PBMC according to gating proposed by Marimuthu et al (29) to minimize non-monocyte contamination.

Leukocyte counts from simultaneous routine haematology laboratory testing were used to calculate absolute cell counts.

### 2.3 Gene expression of enriched monocytes

CD14^+^ monocytes were enriched from thawed PBMC using magnetic beads and a negative selection process (Human Monocyte Enrichment Kit without CD16 Depletion, Stemcell Technologies, Grenoble, France). Post-enrichment monocyte purity was assessed by flow cytometry (mean [range] post-enrichment purity 83.5% [67.5% - 94.5%]). Pre- and post-isolation monocytic HLA-DR MFI remained strongly correlated (r^2^ 0.88, p<0.001) and did not change significantly as a result of enrichment (mean [95% CI] MFI change: -300 [-686 – 86], *P* 0.12).

Total RNA was extracted from enriched monocytes by column-based isolation (RNEasy Micro Kit, Qiagen, Manchester, United Kingdom). 25ng RNA was hybridized to fluorescent barcode-labeled probes from the nCounter Myeloid Innate Immunity V2 Panel (Reference: XT_PGX_HuV2_Myeloid_CSO, Nanostring Technologies, Seattle, United States) prior to analysis on the nCounter SPRINT platform (Nanostring, Technologies, Seattle, United States).

Count data was initially analyzed by ROSALIND (https://rosalind.bio/), with a HyperScale architecture developed by ROSALIND, Inc. (San Diego, CA). Normalization, fold changes and p-values were calculated using criteria provided by Nanostring, following the nCounter Advanced Analysis protocol of dividing counts within a lane by the geometric mean of the normalizer probes from the same lane. Housekeeping probes to be used for normalization were selected based on the geNorm algorithm as implemented in the NormqPCR R library (30). Fold changes and pValues are calculated using the fast method as described in the nCounter® Advanced Analysis 2.0 User Manual.

P-value adjustment is performed using the Benjamini-Hochberg method of estimating false discovery rates (FDR). Downstream analysis was performed using R 4.1.1 and the *ClusterProfiler 4*.*0*.*5, fgsea 1*.*18*.*0* and *gplots 3*.*1*.*1* packages.

### 2.4 Cytokine bead array

Cytokines in thawed serum collected at trough immunosuppression and in parallel with collection of blood for leucocyte profile were quantified by bead-based immunoassay in a representative subset of RTR (n=40), using the ‘LegendPLEX Human Essential Immune Response Panel’ according to manufacturer’s instructions. This panel simultaneously quantifies 13 cytokines essential for immune response: IL-4, IL-2, CXCL10 (IP-10), IL-1β, TNF-α, CCL2 (MCP-1), IL-17A, IL-6, IL-10, IFN-γ, IL-12p70, CXCL8 (IL-8), and Free Active TGF-β1. Results below the limit of detection were accounted for in two ways; firstly analysis was performed with an imputed value 0.1pg/mL below the limit of detection. Analysis was then repeated with exclusion of these samples (the second approach did not change the significance of results and thus is not reported).

### 2.5 Statistical tests

The primary clinical outcome of the study was time from enrolment to first diagnosis of malignancy during follow-up. Diagnoses of cutaneous malignancy were made histologically by trained dermatopathologists: where diagnostic uncertainty was present, the pathologist’s decision was considered final. If the histologist was unable to provide a favoured diagnosis, a non-diagnosis of SCC was presumed. SCC arising within scar tissue or clinically described as recurrence was excluded. Patients were censored at time of date of last follow-up, date of death/graft loss or 365 days, whichever occurred first.

Analyses were performed on Graphpad Prism for Windows 9 (Graphpad Software Inc., San Diego, CA), IBM SPSS 27 and R 4.1.1. Continuous variables are shown as mean±standard deviation (SD) unless specified otherwise., Hazard and odds ratios are reported as ratio (95% confidence interval). Categorical variables are reported as number (percentage of group). For continuous variables, comparison between groups was performed using non-paired two-tailed t-test (two groups) or analysis of variance with post-hoc testing when initial testing was significant (multiple groups) unless specified otherwise. Correlations were tested using Spearman’s test, which is more appropriate for non-parametric data.

Change in mHLA-DRd over time and with malignancy development during follow-up was assessed by mixed-effects modelling. Receiver-operator characteristic (ROC) curves were generated using the *pROC* 1.18.0 package. Hazard ratios (HR) for cancer development during follow-up were generated by creating two models using the *cmprisk* and *finalfit* packages. Firstly, Cox proportional hazards modelling was used to create univariable and multivariable models. Secondly, competing-risks survival regression, based on Fine and Gray’s proportional subhazards model, was undertaken to model for competing risk (graft loss and death). Multivariate regression was undertaken using simultaneous entry into the model, using variables identified as significant upon univariate analysis alongside mHLA-DRd tertile and prednisolone use.

Throughout the study, a *P* value of less than 0.05 was considered statistically significant. Additional supporting information may be found online in the Supporting Information section at the end of the article.

## 3 Results

137 RTR with stable transplant function and no history of recent non-keratinocyte malignancy were recruited, of which 135 had HLA-DR binding density assessed (method demonstrated in Figure 1). Only those with mHLA-DRd quantification were included in subsequent analyses. All non-immunosuppressed controls (n=29) had HLA-DR binding density assessed.

### 3.1 Cohort demographics

The demographics of the cohort are summarised in Table 1. Most participants had a cumulative immunosuppression duration of over twenty years, received no induction therapy at time of transplant and were receiving calcineurin inhibitor-based immunosuppression. The majority were on dual immunosuppression. Non-immunosuppressed controls were around a decade older and had better renal function.

**Table 1:** Cohort characteristics at recruitment. Categorical values are provided as n (%) whilst continuous variables are reported as mean±SD. ‘NA’ not assessed/applicable. Data was available for all participants.

| Characteristic | Transplant Recipients<br>(n=135) | Controls (n=29) |
| --- | --- | --- |
| Male gender | 92 (68.1) | 16 (55.2) |
| Caucasian ethnicity | 131 (97.0) | 29 (100) |
| Age (years) | 62.6±10.8 | 74.9±8.9 |
| >1 renal transplant | 25 (18.5) | NA |
| Body mass index (kg/m<sup>2</sup>) | 25.8±4.7 | NA |
| Previous or current smoker | 52 (38.5) | NA |
| Cytomegalovirus (CMV) seropositive | 86 (63.7) | NA |
| Total HLA mismatch | 2.6±1.5 | NA |
| Total duration of immunosuppression<br>(months) | 244±102 | NA |
| Time since most recent transplant<br>(months) | 223±99 | NA |
| eGFR at recruitment (mL/min/1.73m<sup>2</sup>) | 48.8±19.1 | 70.0±19.1 |
| Induction therapy at most recent<br>transplant |  | NA |
| None | 99 (73.3) |  |
| Thymoglobulin | 7 (5.2) |  |
| Alemtuzumab | 10 (7.4) |  |
| Basiliximab | 19 (14.1) |  |
| Immunosuppression use at recruitment |  | NA |
| Calcineurin inhibitor | 107 (79.3) |  |
| Azathioprine | 89 (65.9) |  |
| Mycophenolate mofetil | 22 (16.3) |  |
| Prednisolone | 46 (34.1) |  |
| Sirolimus | 13 (9.6) |  |
| Number of immunosuppressive agents at<br>recruitment |  | NA |
| 1 | 17 (12.4) |  |
| 2 | 98 (71.5) |  |
| 3 | 22 (16.1) |  |
| History of previous non-keratinocyte<br>malignancy | 16 (11.9) | NA |
| History of previous SCC | 59 (43.7) | 20 (68.9) |

### 3.2 mHLA-DRd is reduced in transplant recipients taking corticosteroids, compared to non-immunosuppressed controls

mHLA-DRd was approximately normally distributed with a degree of positive skew across the transplant cohort (Figure 2). The vast majority of RTR demonstrated mHLA-DRd within the range published for healthy controls, without difference between sex (>15000 antibodies (Ab)/cell, Figure 2) (25). Two participants demonstrated mHLA-DRd within the previously defined immunodepressed range (25) (8000-15000 Ab/cell) and none within the immunoparesis range (<8000 Ab/cell). Mean mHLA-DRd was reduced by nearly 20% in the transplant recipients compared to the non-immunosuppressed control population (40226±2503 vs 32570±1084 Ab/cell, *p*=0.004).

**Figure 2:**
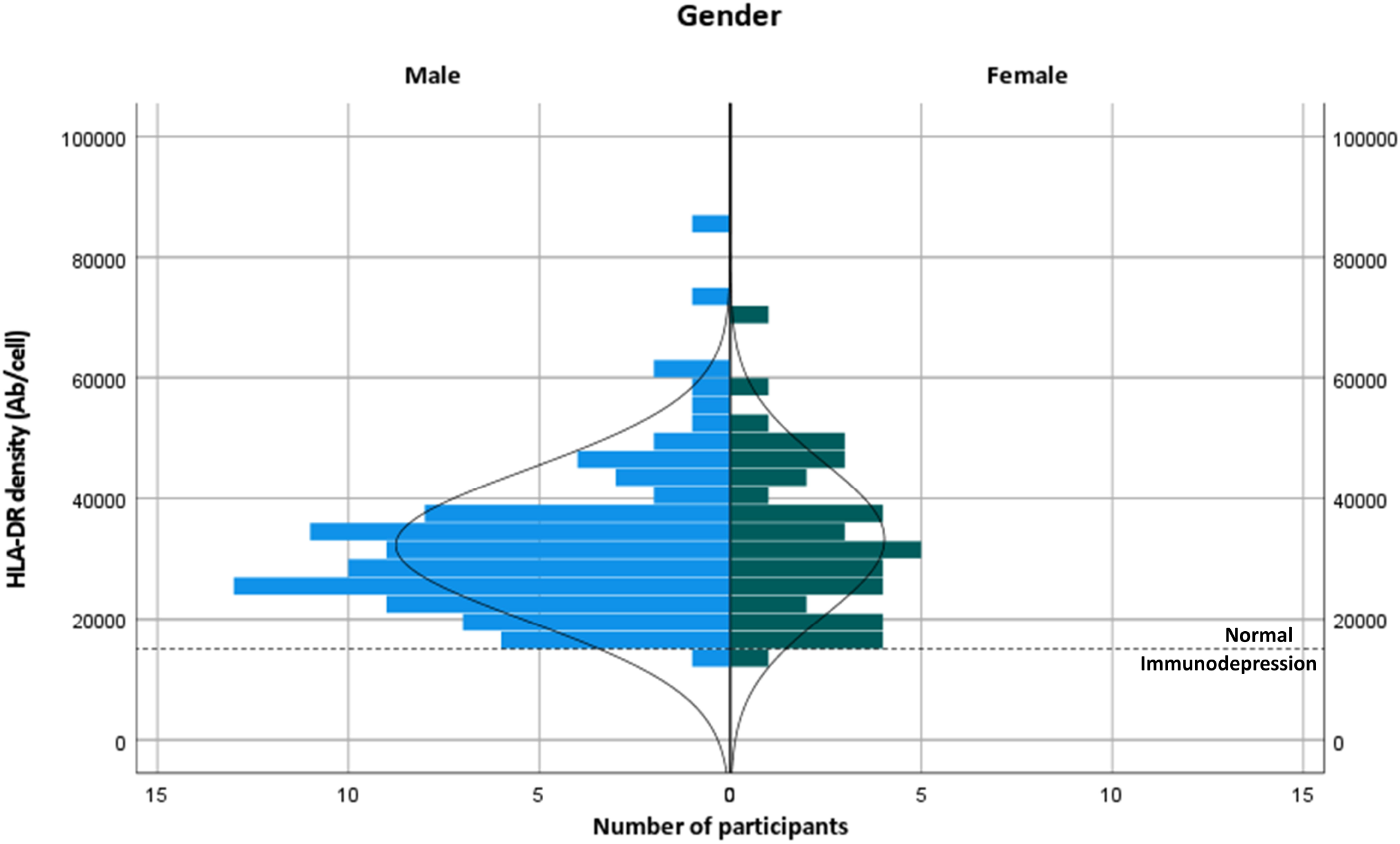
Distribution of HLA-DR binding density in transplant recipients, stratified by gender. The broken line indicates the cut-off for diagnosis of immunodepression as described in (25).

We next asked what factors at recruitment might influence mHLA-DRd in otherwise stable transplant recipients (Figure 3 and Table 2). On univariate analysis after removal of outliers (n=3), no major demographic factor was associated with mHLA-DRd. Notably, there was a significant reduction in mHLA-DRd in RTR receiving corticosteroid therapy and a stepwise and significant decrease in mHLA-DRd by number of agents taken, though a significant difference was only seen between RTR receiving monotherapy and triple therapy on post-hoc testing. RTR on dual and triple therapy exhibited significantly lower mHLA-DRd compared to non-immunosuppressed controls.

**Figure 3:**
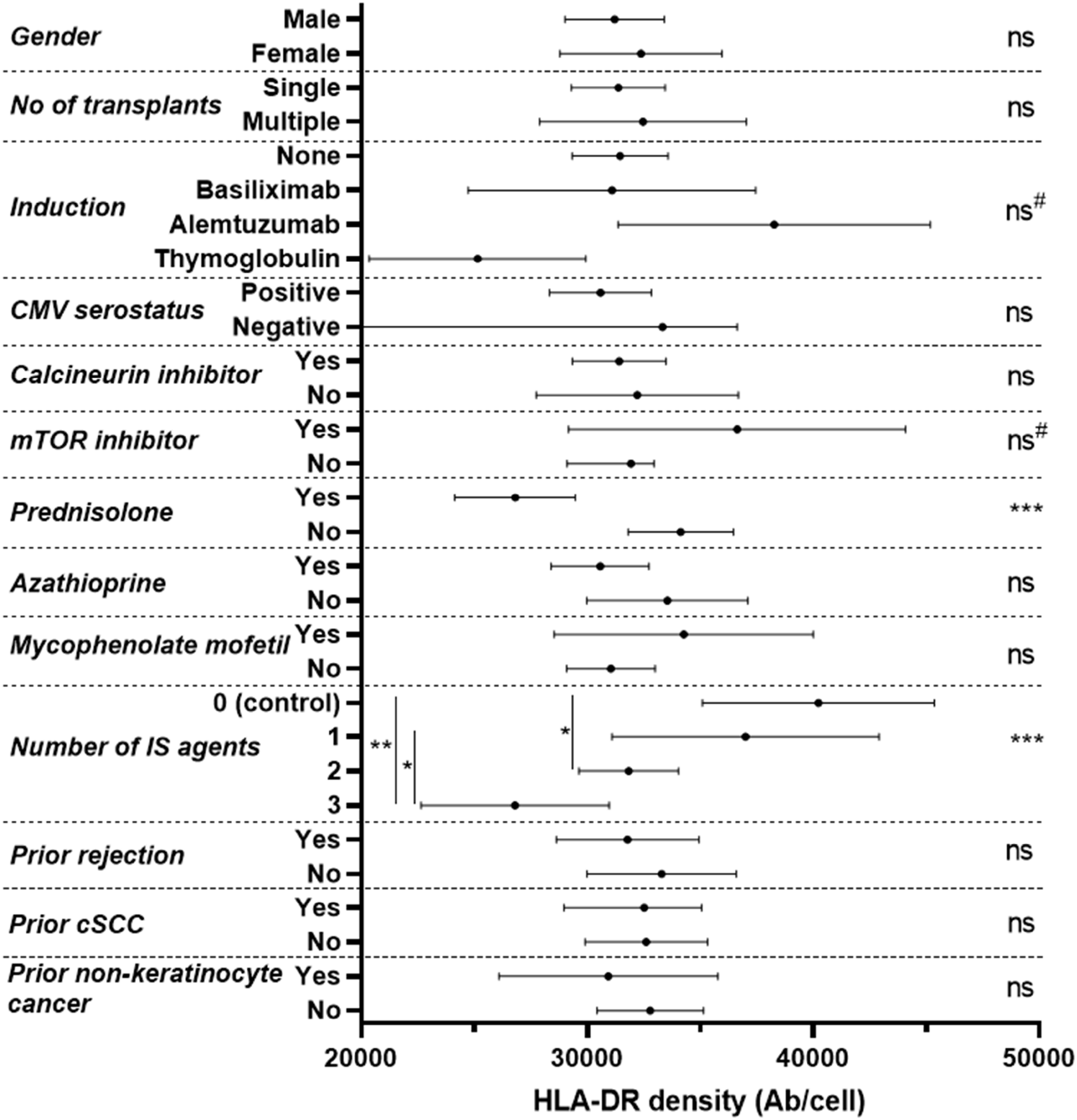
Forest plot of discrete variables at recruitment and their univariable influence upon mHLA-DRd in renal transplant recipients. Densities are recorded as mean and 95% confidence interval, with p values assessed by two-tailed independent t-test or analysis of variance with post-hoc Tukey testing. ‘ns’ not significant; * p<0.05; **p<0.01; ***p<0.001 across all groups (ANOVA or t-test, indicated on right, with post-hoc testing, where appropriate, indicated on left with the vertical line indicating comparison groups). ^#^p<0.10.

**Table 2:** Univariate analysis of continuous variables and their influence upon mHLA-DRd in renal transplant recipients. Spearman’s correlation is reported (r) and significance of correlation.

| Factor | Univariate Correlation | P |
| --- | --- | --- |
| Age | -0.02 | 0.80 |
| Body Mass Index | -0.02 | 0.85 |
| Total Immunosuppression Duration | -0.08 | 0.39 |
| Time since last transplant | -0.10 | 0.26 |
| Total HLA mismatch | -0.08 | 0.38 |
| eGFR at recruitment | 0.07 | 0.42 |
| Serum C-reactive protein within 14 days of recruitment | -0.21 | 0.14 |

Due to the potential for overlapping and conflicting influence of immunomodulatory agents, we undertook a multivariate regression analysis incorporating induction therapy, use of sirolimus or prednisolone at recruitment and number of immunosuppressive agents at recruitment as covariates.

Age at recruitment was also included as a pre-specified variable, as immune ageing can lead to chronic innate immune activation and has recently been found to associate with alterations in monocyte function in renal transplant recipients.^31,32^

This model demonstrated that prednisolone use at recruitment independently predicted a reduction in mHLA-DRd (Beta = -0.30, t(126) = -2.86, p=0.005) with a trend towards the opposite in RTR taking sirolimus at recruitment (Beta = 0.16, t(126) = 1.89, p=0.06). It should be noted that the magnitudes of these effects were relatively small. Induction agent and number of immunosuppressive agents taken did not remain predictive after adjustment (Supplementary Table 2).

There was no clear dose-response relationship with regard to corticosteroid therapy – whilst all RTR taking prednisolone demonstrated reduced mHLA-DRd compared to those not taking it, this did not appear to correlate with weight-adjusted dose taken (Supplementary Figure 2).

### 3.3 mHLA-DRd remains stable during follow-up

We next assessed the medium-term stability of mHLA-DRd. RTR were re-assessed after an interval of a median (IQR) of 238 (196 – 250, n=116) and 385 (264 – 434, n=91) days. Repeat mHLA-DRd exhibited a stable moderate to strong correlation compared to enrolment values (Figure 4) and did not demonstrate a significant shift from baseline values over study follow-up (F(1.86, 189.1) = 0.43, p=0.64). 11 participants had a reduction in the dose of one or more of their immunosuppressive agents during the study follow-up period with mHLA-DRd quantified before and after; no significant change between these two timepoints was seen (data not shown).

**Figure 4:**
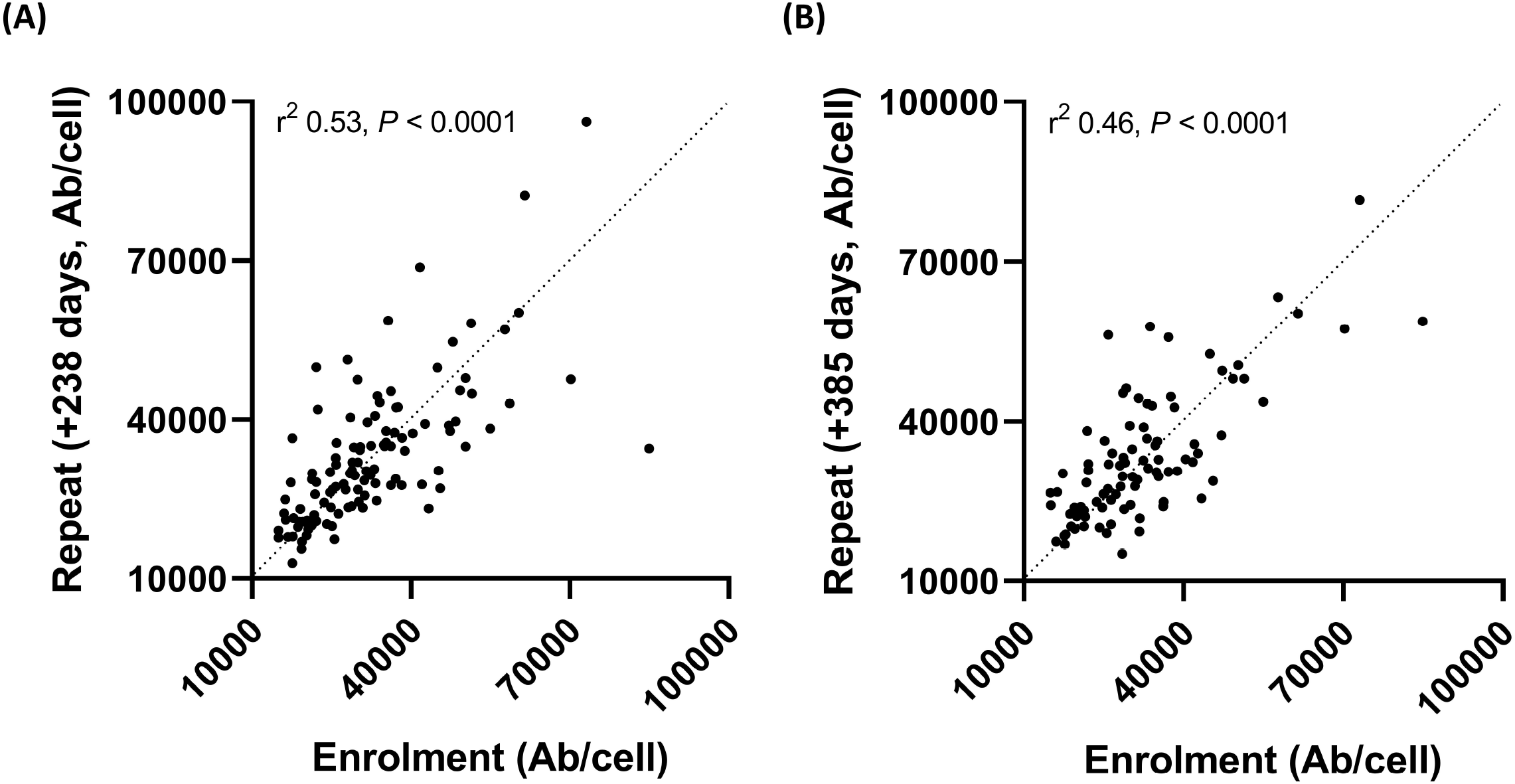
Monocytic HLA-DR density shows stability over time. mHLA-DRd was calculated at enrolment and repeated a mean of 238 (a) and 385 (b) days later. Spearman’s test was used to assess goodness-of-fit (r^2^) and significance.

### 3.4 mHLA-DRd is independent of costimulatory receptor density and correlates with the accumulation of monocytes with an MDSC-like phenotype

We next assessed correlation between mHLA-DRd and lymphocyte and monocyte populations.

The total monocyte number in transplant recipients demonstrated no correlation with mHLA-DRd (r -0.11, p 0.20). Similarly, no correlation was found with total lymphocyte count or with major T cell subsets (r <0.2 for all populations studied, Supplementary Figure 3). Specifically, we previously reported an increased risk of cutaneous malignancy in transplant recipients exhibiting evidence of T cell immunosenescence (28) – no correlation was seen between mHLA-DRd and phenotypic markers of this. In those not taking azathioprine, which has a marked effect upon B and NK cell populations (28), no correlation was seen with overall number or subpopulation proportion (Supplementary Figure 3).

A subset of patients (n=14) at the extremes of mHLA-DRd underwent more detailed characterization of the monocyte compartment on thawed PBMC (Figure 5A). mHLA-DRd showed weak and no correlation with overall expression of the costimulatory receptors CD86 and CD80 upon monocytes respectively (Supplementary Figure 4A). Decreasing mHLA-DRd was moderately associated with a increase in the proportion of CD14^+^CD16^lo^ classical and a reduction in the proportion of non-classical CD14^lo^CD16^+^ subsets, with no change in the proportion of intermediate monocytes (Supplementary Figure 4B). However, mHLA-DRd correlated strongly with the accumulation of CD19^-^CD3^-^CD56^-^CD14^+/int^CD11b^+^CD33^+^HLA-DR^lo^ myeloid-derived suppressor cell-like monocytes (mMDSC-like), both in terms of calculated absolute number and proportion of the total monocyte gate (Figure 6B and 6C). Goodness-of-fit (r^2^) testing suggested mMDSC-like cell accumulation explained around 60-65% of mHLA-DRd variation at a univariate level. There was no difference between monocyte populations when stratified by corticosteroid use (Supplementary Figure 4C).

**Figure 5:**
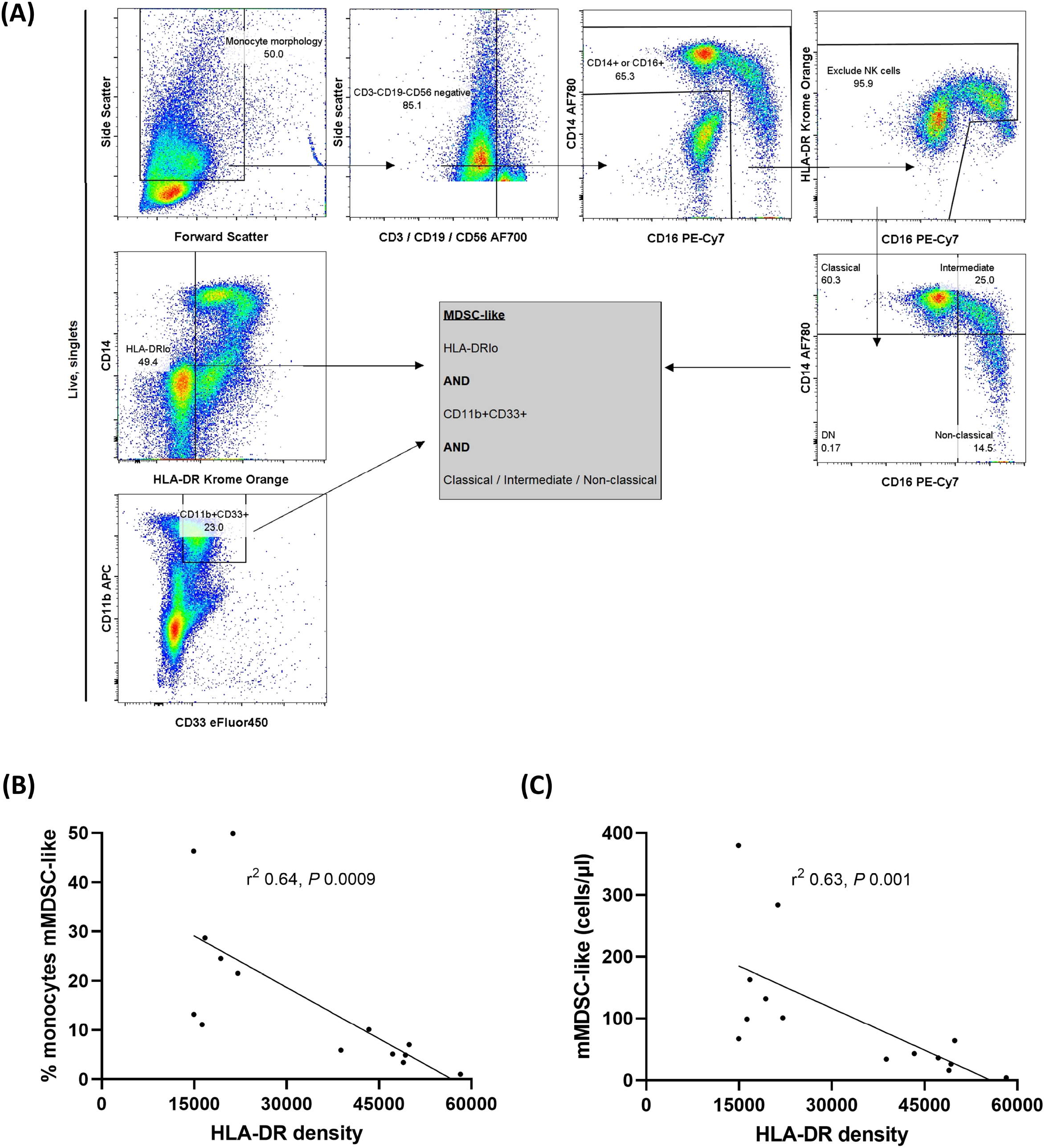
mHLA-DRd correlates inversely with CD19^-^CD3^-^CD56^-^CD14^+/int^CD11b^+^CD33^+^HLA-DR^lo^ monocyte myeloid-derived suppressor-like cell accumulation. (A) Example of gating strategy to delineate monocyte subpopulations. Correlation between mHLA-DRd and (B) proportion of mMDSC-like cells within the monocyte population and (C) absolute number of mMDSC-like cells. Spearman’s test was used to assess goodness-of-fit (r^2^) and significance.

**Figure 6:**
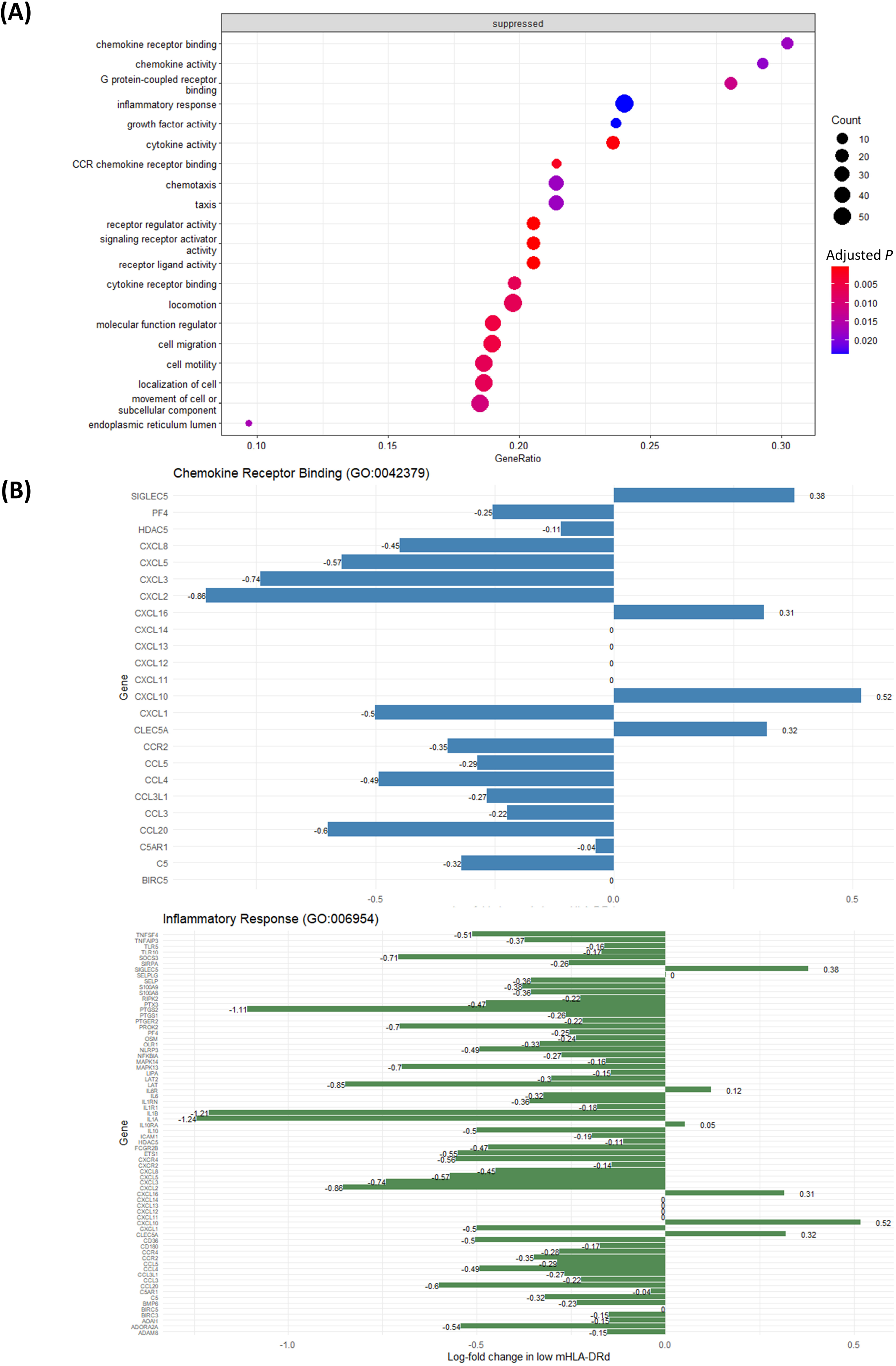
Monocytes from RTR with low mHLA-DRd exhibit suppression of gene sets relating to inflammatory response and chemotaxis. (A) Pathways with significantly altered enrichment in RTR exhibiting low mHLA-DRd, using the Gene Ontology knowledge base. Pathways are listed in order of descending GeneRatio. (B) Differential expression of core genes in ‘Chemokine receptor binding’ and ‘Inflammatory response’ Gene Ontology sets.

### 3.5 Reduced mHLA-DRd is associated with dampening of inflammatory signalling pathways in the absence of major changes in circulatory cytokine concentration

In order to assess what transcriptomic pathways might be driving the changes in mHLA-DRd across the monocyte population, total RNA from enriched monocytes from RTR with high and low levels of mHLA-DRd, matched for age and prednisolone use, was analysed using the nCounter ‘Human Myeloid Innate Immunity’ panel. This panel includes 770 genes in 19 different pathways and processes related to myeloid cell function. The details of participants included in this analysis are given in Supplementary Table 3.

Surprisingly, unsupervised principle component analysis of did not suggest major transcriptomic differences at a whole panel level, based on prednisolone use or mHLA-DRd (Supplementary Figure 4A). Similarly, differential expression analysis revealed no major changes in gene expression based on mHLA-DRd when adjusted for multiple testing, though a number were differentially expressed on unadjusted testing (Supplementary Figure 4B). A subset of genes associated with mMDSC were assessed specifically (31); these did not cluster based on mHLA-DRd (Supplementary Figure 4C).

To assess for smaller but coordinated perturbations within pathways not visible upon differential expression analysis, gene set enrichment analysis was performed using the Gene Ontology molecular functions and biological processes knowledge bases (32). This revealed suppression of gene sets relating to inflammation, cytokine and chemokine signalling and chemotaxis and cellular locomotion in monocytes with low mHLA-DRd (Figure 6A and Supplementary Table 4); upon interrogation of these sets, widespread downregulation of Toll-like receptor, chemokine and cytokine ligand and receptor expression, as well as their downstream mediators, was noted (Figure 6B).

Given the suppression of gene sets relating to inflammation and cytokine signalling in monocytes with low mHLA-DRd, we questioned whether this was indicative of evidence of reduced systemic inflammation. Serum c-reactive protein (CRP) and 13 cytokines were assessed. In those with a serum C-reactive protein (CRP) measurement within 14 days of sampling (n=53, where 45 were taken simultaneously with immune profiling), there was no correlation between CRP concentration and mHLA-DRd (data not shown). Similarly, whilst all cytokines demonstrated strong correlation with each other, none demonstrated significant correlation with mHLA-DRd (Supplementary Figure 6). There was no difference in the serum concentration of any cytokine when stratified by prednisolone use (data not shown).

Taken together, the above results suggest that decreased mHLA-DRd is associated with dampened inflammatory signalling and reduced chemotactic potential within the circulating monocyte pool, in the absence of a discernable change in serum cytokines and markers of systemic inflammation.

### 3.6 Reduced mHLA-DRd associates with increased subsequent malignancy risk

Given literature suggesting mHLA-DRd may be a biomarker of ‘over-immunosuppression’ and the above data suggesting accumulation of MDSC-like cells and reduced inflammatory and chemotactic response in monocytes from RTR with low mHLA-DRd, we hypothesised that chronic reduction of mHLA-DRd would associate with other complications associated with immunosuppression, namely malignancy. Outcomes in the first 365 days after sampling were analysed, i.e. during the period where mHLA-DRd stability had been demonstrated. During this period, three participants were lost to follow-up (one loss to follow-up, one each due to death and graft loss). 28 RTR (21%) developed malignancy, of which 25 represented cutaneous SCC as the first malignancy during follow-up. The mean±SD time to malignancy diagnosis after recruitment was 190±114 days.

Patients who developed malignancy during follow-up trended towards lower mHLA-DRd on serial sampling than those who did not, though there was significant overlap (F(1, 133) = 2.06, P = 0.15, Supplementary Figure 7). A receiver-operator characteristic (ROC) curve was generated which suggested a cut-off of 29912 would generate an area under the curve (AUC) of 0.635, representing a sensitivity of 0.58 and specificity of 0.68 for malignancy development within the following year (Figure 7A). Univariable analysis confirmed those with a mHLA-DRd below this cut-off were at double the risk of subsequent malignancy during the following year (Figure 7B and Table 3). This propensity remained significant upon multivariable adjustment and upon correcting for the competing risk of death or graft loss (Table 3).

**Figure 7:**
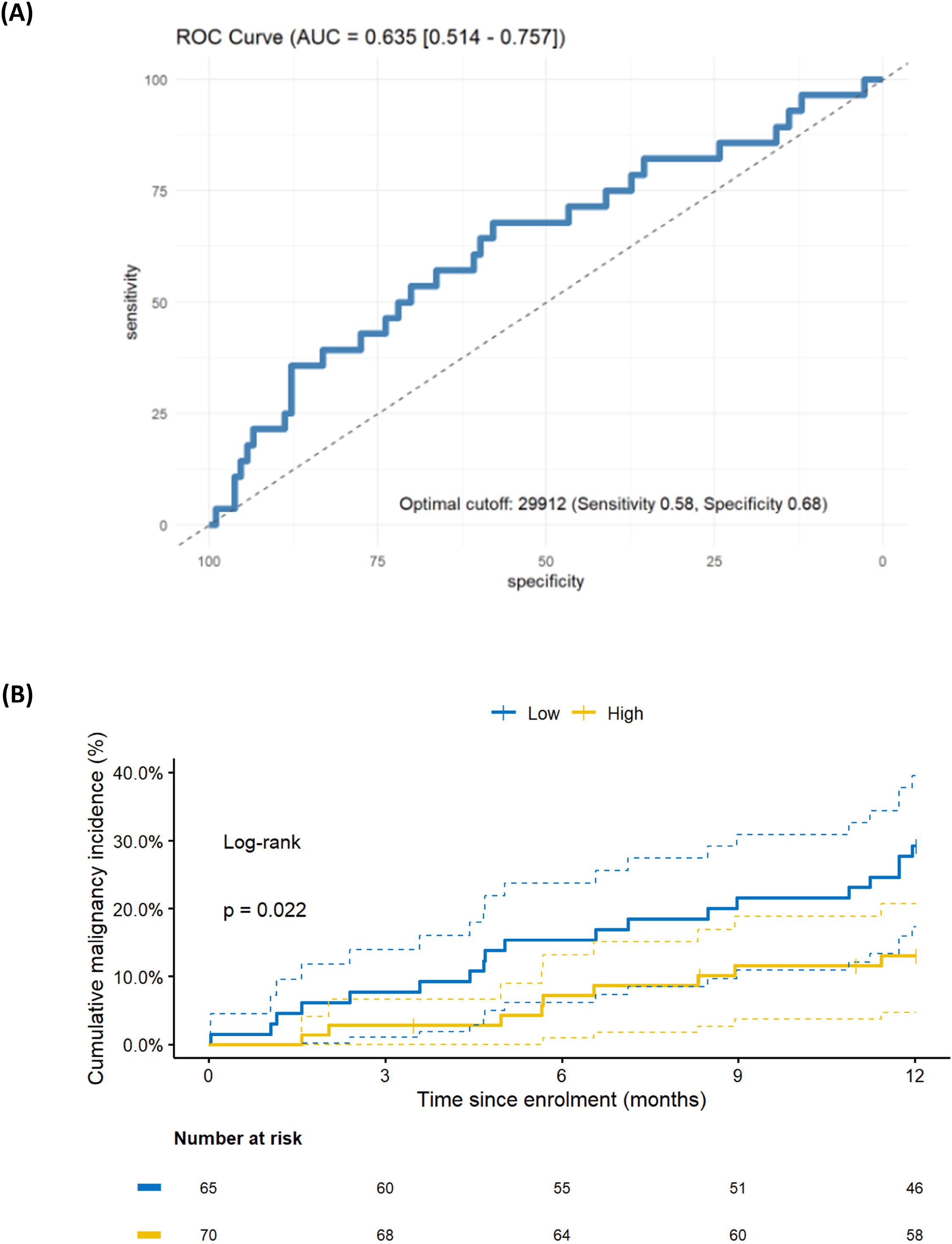
Decreased mHLA-DRd is associated with subsequent malignancy development. (A) Receiver-operator characteristic (ROC) curve for prediction of malignancy in the year following mHLA-DRd quantification. Predictive performance is given (area under curve (AUC) and 95% confidence interval) as well as the optimal cut-off and corresponding sensitivity and specificity. (B) Kaplan-Meier curve demonstrating cumulative incidence of malignancy stratified by mHLA-DRd. (unadjusted and adjusted hazard ratios and *P* values are given in Table 3; log-rank test for difference between curves is provided in-plot).

**Table 3:**
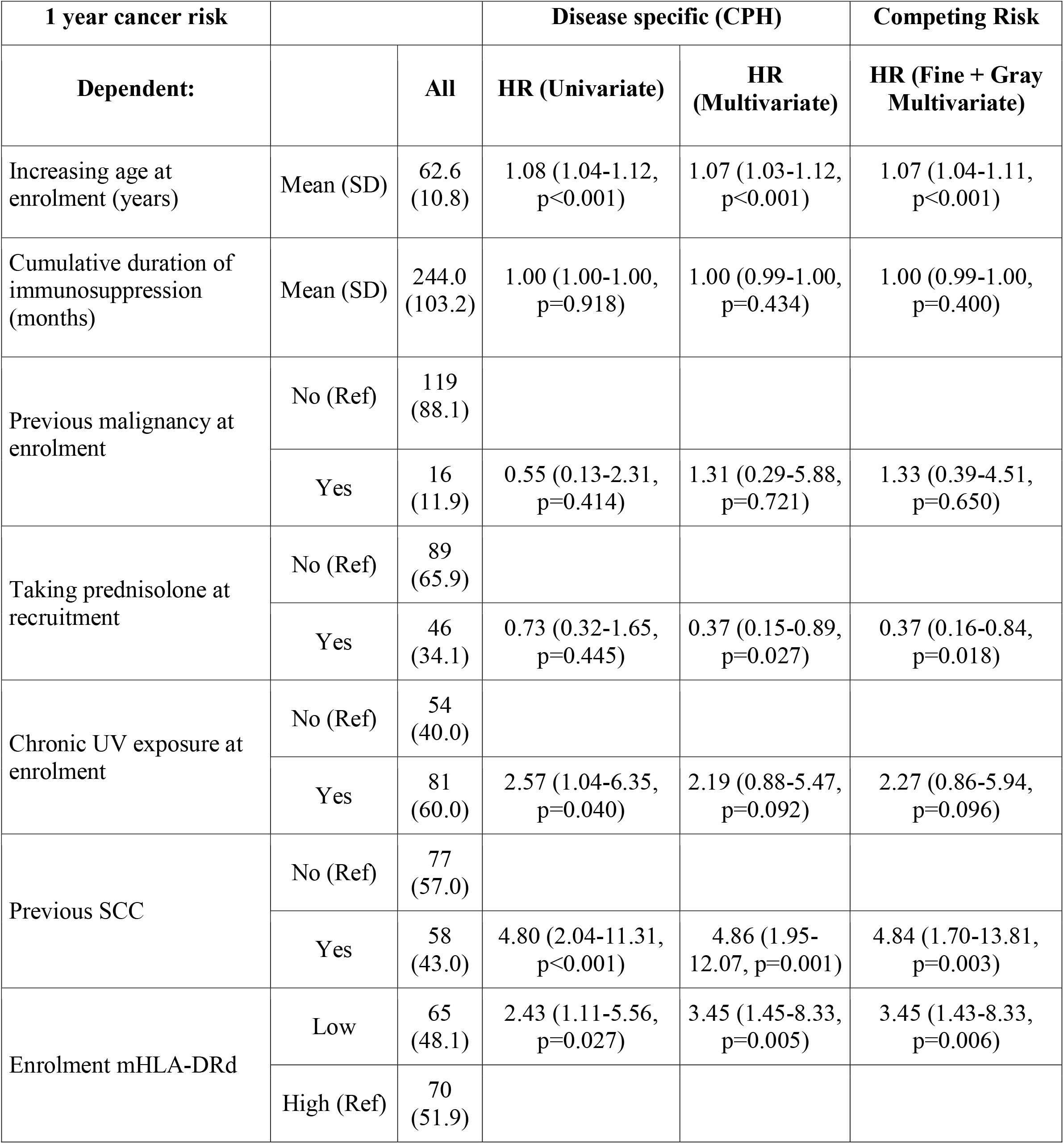
Hazard ratios for malignancy development within one year of mHLA-DRd quantification. All factors analysed for univariate significant were included in multivariate models as a pre-specified analysis. Age at recruitment and age at last transplant demonstrated strong correlation, as did number of immunosuppressive agents and the use of prednisolone at recruitment, and so only age and use of prednisolone at recruitment were used in the multivariate models. Two models were generated: one using Cox Proportionate Hazards modelling and a second using competing-risk modelling, using graft loss and death as competing risks.

## 4 Discussion

Malignancy is a major contributor to patient morbidity and mortality, which remains relatively unchanged over the last thirty years (9). Greater understanding of the mechanism which predispose to cancer development are sorely needed, to facilitate studies into targeted intervention. Based on data from short-term outcomes in lung and liver transplantation, with renewed interest in mHLA-DRd driven by trials of cell therapy, we investigated the mechanisms and consequences of reduced mHLA-DRd in long-term transplant outcomes. The use of a long-term population at high risk of malignancy development, by virtue of their cumulative immunosuppressive burden or previous keratinocyte malignancy, enriched our study population for the outcome of interest (namely cancer), allowing easier study of contributory factors.

We found that mHLA-DRd is a distinct immunological parameter, demonstrating stability over a twelve month period, that does not correlate with clinically utilised inflammatory markers, major circulating cytokines, or circulating lymphocytes, but appears to reflect accumulation of CD14^+^CD11b^+^CD33^+^HLA-DR^lo^ cells within the monocyte compartment. Monocytic myeloid-derived suppressor cells (mMDSC) are a heterogenous population and a definitive marker is yet to be described (31, 33), but cells with this phenotype have been previously demonstrated to suppress T and NK cell proliferation *in vitro* through a number of mechanisms, including induction of regulatory T cells, metabolite depletion, and secretion of reactive oxygen species (ROS) and nitric oxide (NO) (34, 35). Recent work in mice comparing MSDC populations and transcriptional pathways in the setting of chronic viral infection or malignancy suggests different mechanisms may predominate depending on the context of the stimulus (36). It has been suggested that cells with a MDSC phenotype but where suppressive function has not been demonstrated should be termed (monocytic) ‘myeloid derived suppressor-like cells’ ((m)MDS-LC) (31).

When we investigated transcriptomic changes that underlie the reduction in mHLA-DRd, we did not find any significantly differentially expressed genes after correction for multiple testing. There are three possible explanations for this: the first is that this was an assessment of the whole monocyte compartment and thus bulk RNA analysis may miss subtle changes within one subset (i.e. accumulation of mMDS-LC, as evidenced by a lack of clustering of MDSC-related genes). Secondly, the rapid upregulation of mHLA-DRd *ex vivo* when stored at room temperature (Supplementary Figure S1) suggests that pathways that control mHLA-DRd in non-suppressive monocyte populations may be regulated at the post-transcriptomic level. The use of gene set enrichment analysis allows for interrogation and identification of pathways that may be involved in this process without assumption about the magnitude of change or use of pre-defined limits. Interrogating the Gene Ontology knowledgebase revealed that monocytes from RTR with low mHLA-DRd exhibited evidence of dampened chemokine and cytokine signalling and chemotaxis. Our data may provide a second, more subtle, mechanism by which reduced mHLA-DRd is associated with poorer outcomes in a number of clinical scenarios, due to dampened responses to cytokine and chemotactic signals leading to altered trafficking of monocytes to the periphery. The lack of correlation with circulating cytokines and CRP suggests that the dampened responses are not due to a systemic reduction in inflammatory mediators.

The effect of corticosteroids upon mHLA-DRd was notable. The suppression of mHLA-DRd was not due to accumulation of MDS-LC, as no major differences were seen in subpopulations. The effect of prednisolone on mHLA-DRd is consistent with other reports but, in contrast to these, we did not see a major difference in monocyte subpopulations (particularly CD14^hi^CD16^-^, where many MDS-LC are found) (22, 37). It should be remarked that the majority of our cohort were on a relatively small daily dose of prednisolone and this might explain the discrepancy in findings. It is likely that corticosteroids dampen inflammatory response pathways, leading to decreased mHLA-DRd, independent of the effect upon monocyte populations or circulating markers of inflammation.

Given the association with accumulation of mMDS-LC and evidence of reduced inflammatory response, we sought to assess whether reduced mHLA-DRd was associated with poorer medium-term outcomes; namely increased risk of subsequent malignancy, even though the vast majority of RTR demonstrated mHLA-DRd within the previously cited ‘normal’ range (25). The monocyte-macrophage axis plays a complex role in cancer development, with evidence of both pro- and anti-tumour functions (38, 39). Circulating m-MDSC accumulate early after renal transplant and may identify RTR at risk of early post-transplant cancer (40, 41). m-MDSC numbers were highly variable during the early post-transplant period and whether predictive value remained beyond this timeframe is unclear. m-MDSC expansion may also promote regulatory T cell (Treg) expansion *in vitro* and in animal models (40); notably we did not observe a correlation between circulating Treg and reduced mHLA-DRd, though this does not preclude peripheral expansion of Treg. Cutaneous SCC was the most common cancer seen during follow-up and it is noteworthy that mMDSC are implicated both in the mutagenesis and progression of cSCC. mMDSC are induced by some strains of beta-genus human papillomavirus, the presence of which has been controversially linked to cutaneous SCC development (38, 42, 43). In mouse model of UV radiation and HPV oncogene-induced cSCC, Ly6C^hi^ monocytes infiltrate and accumulate at the site of UV radiation injury and are essential for carcinogenesis (44); however in this model the mechanism appears to be through their pro-inflammatory effects. The Th2-polarised cytokine state seen within cutaneous SCC from transplant recipients may lead to mMDSC induction and reduced circulating mHLA-DRd (38, 45). The potential use of mHLA-DRd as a marker of mMDSC accumulation is appealing in that standard curve generation and the simplicity of gating is likely to be more reproducible in multi-centre studies and in clinical practice (25). How the changes in circulating monocyte phenotype relates to changes in peripheral monocyte behaviour are a focus of ongoing study.

Whilst this study provides important insight into the clinical factors and mechanisms that drive mHLA-DRd and the clinical sequelae of reductions in this, there were some limitations. Survivor bias of long-term RTR at recruitment may diminish the apparent association between mHLA-DRd and malignancy. Secondly, cutaneous squamous cell carcinoma was the main malignancy type in this study, reflecting its frequency amongst long-term, Caucasian RTR (2). Whether reduced mHLA-DRd predicts other malignancy in other ethnic populations or in other settings where immunosuppression is used, such as autoimmunity, is unclear. Due to the exploratory nature of this study, no *a priori* power calculation was performed and this leaves a possibility of a type I error; however our data would be consistent with, though an extrapolation of, findings demonstrating an impaired immune response with diminished mHLA-DRd in multiple other settings. Finally, only a small subset of our cohort had a change in their immunosuppressive therapy during study follow-up, and these changes were highly heterogeneous; therefore the extent to which mHLA-DRd is dynamic as a response to changes in immunosuppression is unclear.

In conclusion, we report that reduced mHLA-DRd is a distinct immunological marker that reflects the accumulation of mMDSC-like cells within the monocyte population and dampening of pathways relating to inflammation, cytokine signalling and chemotaxis. This is clinically relevant in that it is associated with the subsequent development of malignancy. Our findings will drive further study into the mechanisms underlying this association with cancer development, which may reveal novel avenues for therapeutic manipulation.

## Supporting information

checklist

## Data Availability

All data produced in the present study are available upon reasonable request to the authors.

## 6 Author Contributions

The study was devised by MJB, KJW and PNH. Recruitment and data collection was undertaken by MJB. Analysis and synthesis of conclusions were undertaken by all authors. Manuscript drafting was undertaken by MJB, with revision and comments by JH, FI, PNH and KJW.

## 8 Acknowledgments

The authors would like to thank the staff and patients at the Oxford Transplant Unit for their support of this study, in particular S. Ruse for assisting in coordinating the follow-up of participants. We are grateful to G. Betts and R. Arroyo-Hornero for their technical assistance and A. Cross for her review of the manuscript.

The authors acknowledge the support of the National Institute for Health Research, through the Local Clinical Research Network.

## 9 Data Availability Statement

Original datasets will be deposited in a publicly accessible repository prior to final publication. In the interim, the datasets are available from the corresponding author upon reasonable request.

## References

1. Ying T, Shi B, Kelly PJ, Pilmore H, Clayton PA, Chadban SJ. Death after Kidney Transplantation: An Analysis by Era and Time Post-Transplant. J Am Soc Nephrol (2020) 31(12):2887–99. Epub 20200909. doi: 10.1681/ASN.2020050566.

2. Hartevelt MM, Bavinck JN, Kootte AM, Vermeer BJ, Vandenbroucke JP. Incidence of Skin Cancer after Renal Transplantation in the Netherlands. Transplantation (1990) 49(3):506–9. doi: 10.1097/00007890-199003000-00006.

3. Harwood CA, Mesher D, McGregor JM, Mitchell L, Leedham-Green M, Raftery M, et al. A Surveillance Model for Skin Cancer in Organ Transplant Recipients: A 22-Year Prospective Study in an Ethnically Diverse Population. Am J Transplant (2013) 13(1):119–29. Epub 20121016. doi: 10.1111/j.1600-6143.2012.04292.x.

4. Rosales BM, De La Mata N, Vajdic CM, Kelly PJ, Wyburn K, Webster AC. Cancer Mortality in Kidney Transplant Recipients: An Australian and New Zealand Population-Based Cohort Study, 1980-2013. Int J Cancer (2020) 146(10):2703–11. Epub 20190823. doi: 10.1002/ijc.32585.

5. Vajdic CM, McDonald SP, McCredie MR, van Leeuwen MT, Stewart JH, Law M, et al. Cancer Incidence before and after Kidney Transplantation. JAMA (2006) 296(23):2823–31. doi: 10.1001/jama.296.23.2823.

6. Villeneuve PJ, Schaubel DE, Fenton SS, Shepherd FA, Jiang Y, Mao Y. Cancer Incidence among Canadian Kidney Transplant Recipients. Am J Transplant (2007) 7(4):941–8. Epub 20070228. doi: 10.1111/j.1600-6143.2007.01736.x.

7. Blosser CD, Haber G, Engels EA. Changes in Cancer Incidence and Outcomes among Kidney Transplant Recipients in the United States over a Thirty-Year Period. Kidney Int (2021) 99(6):1430–8. Epub 20201105. doi: 10.1016/j.kint.2020.10.018.

8. D’Arcy ME, Coghill AE, Lynch CF, Koch LA, Li J, Pawlish KS, et al. Survival after a Cancer Diagnosis among Solid Organ Transplant Recipients in the United States. Cancer (2019) 125(6):933–42. Epub 20190109. doi: 10.1002/cncr.31782.

9. Miao Y, Everly JJ, Gross TG, Tevar AD, First MR, Alloway RR, et al. De Novo Cancers Arising in Organ Transplant Recipients Are Associated with Adverse Outcomes Compared with the General Population. Transplantation (2009) 87(9):1347–59. doi: 10.1097/TP.0b013e3181a238f6.

10. Siu JHY, Surendrakumar V, Richards JA, Pettigrew GJ. T Cell Allorecognition Pathways in Solid Organ Transplantation. Front Immunol (2018) 9:2548. Epub 20181105. doi: 10.3389/fimmu.2018.02548.

11. Vester H, Dargatz P, Huber-Wagner S, Biberthaler P, van Griensven M. Hla-Dr Expression on Monocytes Is Decreased in Polytraumatized Patients. Eur J Med Res (2015) 20:84. Epub 20151016. doi: 10.1186/s40001-015-0180-y.

12. Venet F, Monneret G. Advances in the Understanding and Treatment of Sepsis-Induced Immunosuppression. Nat Rev Nephrol (2018) 14(2):121–37. Epub 20171211. doi: 10.1038/nrneph.2017.165.

13. Chenouard A, Rimbert M, Joram N, Braudeau C, Roquilly A, Bourgoin P, et al. Monocytic Human Leukocyte Antigen Dr Expression in Young Infants Undergoing Cardiopulmonary Bypass. Ann Thorac Surg (2021) 111(5):1636–42. Epub 20200708. doi: 10.1016/j.athoracsur.2020.05.071.

14. Haveman JW, van den Berg AP, Verhoeven EL, Nijsten MW, van den Dungen JJ, The HT, et al. Hla-Dr Expression on Monocytes and Systemic Inflammation in Patients with Ruptured Abdominal Aortic Aneurysms. Crit Care (2006) 10(4):R119. doi: 10.1186/cc5017.

15. Sint A, Lutz R, Assenmacher M, Kuchenhoff H, Kuhn F, Faist E, et al. Monocytic Hla-Dr Expression for Prediction of Anastomotic Leak after Colorectal Surgery. J Am Coll Surg (2019) 229(2):200–9. Epub 20190322. doi: 10.1016/j.jamcollsurg.2019.03.010.

16. Zhang R, Shi J, Zhang R, Ni J, Habtezion A, Wang X, et al. Expanded Cd14(Hi)Cd16(-) Immunosuppressive Monocytes Predict Disease Severity in Patients with Acute Pancreatitis. J Immunol (2019) 202(9):2578–84. Epub 20190320. doi: 10.4049/jimmunol.1801194.

17. Benlyamani I, Venet F, Coudereau R, Gossez M, Monneret G. Monocyte Hla-Dr Measurement by Flow Cytometry in Covid-19 Patients: An Interim Review. Cytometry A (2020) 97(12):1217–21. Epub 20201104. doi: 10.1002/cyto.a.24249.

18. Harden PN, Game DS, Sawitzki B, Van der Net JB, Hester J, Bushell A, et al. Feasibility, Long-Term Safety, and Immune Monitoring of Regulatory T Cell Therapy in Living Donor Kidney Transplant Recipients. Am J Transplant (2021) 21(4):1603–11. Epub 20210202. doi: 10.1111/ajt.16395.

19. Roemhild A, Otto NM, Moll G, Abou-El-Enein M, Kaiser D, Bold G, et al. Regulatory T Cells for Minimising Immune Suppression in Kidney Transplantation: Phase I/Iia Clinical Trial. BMJ (2020) 371:m3734. Epub 20201021. doi: 10.1136/bmj.m3734.

20. Sawitzki B, Harden PN, Reinke P, Moreau A, Hutchinson JA, Game DS, et al. Regulatory Cell Therapy in Kidney Transplantation (the One Study): A Harmonised Design and Analysis of Seven Non-Randomised, Single-Arm, Phase 1/2a Trials. Lancet (2020) 395(10237):1627–39. doi: 10.1016/S0140-6736(20)30167-7.

21. Cho JH, Yoon YD, Jang HM, Kwon E, Jung HY, Choi JY, et al. Immunologic Monitoring of T-Lymphocyte Subsets and Hla-Dr-Positive Monocytes in Kidney Transplant Recipients: A Prospective, Observational Cohort Study. Medicine (Baltimore) (2015) 94(44):e1902. doi: 10.1097/MD.0000000000001902.

22. Haveman JW, van den Berg AP, van den Berk JM, Mesander G, Slooff MJ, de Leij LH, et al. Low Hla-Dr Expression on Peripheral Blood Monocytes Predicts Bacterial Sepsis after Liver Transplantation: Relation with Prednisolone Intake. Transpl Infect Dis (1999) 1(3):146–52. doi: 10.1034/j.1399-3062.1999.010302.x.

23. Hoffman JA, Weinberg KI, Azen CG, Horn MV, Dukes L, Starnes VA, et al. Human Leukocyte Antigen-Dr Expression on Peripheral Blood Monocytes and the Risk of Pneumonia in Pediatric Lung Transplant Recipients. Transpl Infect Dis (2004) 6(4):147–55. doi: 10.1111/j.1399-3062.2004.00069.x.

24. Kunz D, Pross M, Konig W, Lippert H, Manger T. Diagnostic Relevance of Procalcitonin, Il-6 and Cellular Immune Status in the Early Phase after Liver Transplantation. Transplant Proc (1998) 30(5):2398–9. doi: 10.1016/s0041-1345(98)00667-8.

25. Docke WD, Hoflich C, Davis KA, Rottgers K, Meisel C, Kiefer P, et al. Monitoring Temporary Immunodepression by Flow Cytometric Measurement of Monocytic Hla-Dr Expression: A Multicenter Standardized Study. Clin Chem (2005) 51(12):2341–7. Epub 20051007. doi: 10.1373/clinchem.2005.052639.

26. von Elm E, Altman DG, Egger M, Pocock SJ, Gotzsche PC, Vandenbroucke JP, et al. Strengthening the Reporting of Observational Studies in Epidemiology (Strobe) Statement: Guidelines for Reporting Observational Studies. BMJ (2007) 335(7624):806–8. doi: 10.1136/bmj.39335.541782.AD.

27. van Vugt MJ, van den Herik-Oudijk IE, van de Winkle JG. Binding of Pe-Cy5 Conjugates to the Human High-Affinity Receptor for Igg (Cd64). Blood (1996) 88(6):2358–61.

28. Bottomley MJ, Harden PN, Wood KJ. Cd8+ Immunosenescence Predicts Post-Transplant Cutaneous Squamous Cell Carcinoma in High-Risk Patients. J Am Soc Nephrol (2016) 27(5):1505–15. Epub 20151112. doi: 10.1681/ASN.2015030250.

29. Marimuthu R, Francis H, Dervish S, Li SCH, Medbury H, Williams H. Characterization of Human Monocyte Subsets by Whole Blood Flow Cytometry Analysis. J Vis Exp (2018) (140). Epub 20181017. doi: 10.3791/57941.

30. Perkins JR, Dawes JM, McMahon SB, Bennett DL, Orengo C, Kohl M. Readqpcr and Normqpcr: R Packages for the Reading, Quality Checking and Normalisation of Rt-Qpcr Quantification Cycle (Cq) Data. BMC Genomics (2012) 13:296. Epub 20120702. doi: 10.1186/1471-2164-13-296.

31. Bronte V, Brandau S, Chen S-H, Colombo MP, Frey AB, Greten TF, et al. Recommendations for Myeloid-Derived Suppressor Cell Nomenclature and Characterization Standards. Nature Communications (2016) 7(1):12150. doi: 10.1038/ncomms12150.

32. Subramanian A, Tamayo P, Mootha VK, Mukherjee S, Ebert BL, Gillette MA, et al. Gene Set Enrichment Analysis: A Knowledge-Based Approach for Interpreting Genome-Wide Expression Profiles. Proc Natl Acad Sci U S A (2005) 102(43):15545–50. Epub 20050930. doi: 10.1073/pnas.0506580102.

33. Veglia F, Sanseviero E, Gabrilovich DI. Myeloid-Derived Suppressor Cells in the Era of Increasing Myeloid Cell Diversity. Nature Reviews Immunology (2021) 21(8):485–98. doi: 10.1038/s41577-020-00490-y.

34. Bruger AM, Dorhoi A, Esendagli G, Barczyk-Kahlert K, Van Der Bruggen P, Lipoldova M, et al. How to Measure the Immunosuppressive Activity of Mdsc: Assays, Problems and Potential Solutions. Cancer Immunology, Immunotherapy (2019) 68(4):631–44. doi: 10.1007/s00262-018-2170-8.

35. Ma T, Renz BW, Ilmer M, Koch D, Yang Y, Werner J, et al. Myeloid-Derived Suppressor Cells in Solid Tumors. Cells (2022) 11(2):310. doi: 10.3390/cells11020310.

36. Tcyganov EN, Hanabuchi S, Hashimoto A, Campbell D, Kar G, Slidel TWF, et al. Distinct Mechanisms Govern Populations of Myeloid-Derived Suppressor Cells in Chronic Viral Infection and Cancer. Journal of Clinical Investigation (2021) 131(16). doi: 10.1172/jci145971.

37. Rogacev KS, Zawada AM, Hundsdorfer J, Achenbach M, Held G, Fliser D, et al. Immunosuppression and Monocyte Subsets. Nephrol Dial Transplant (2015) 30(1):143–53. Epub 20141013. doi: 10.1093/ndt/gfu315.

38. Bottomley MJ, Thomson J, Harwood C, Leigh I. The Role of the Immune System in Cutaneous Squamous Cell Carcinoma. Int J Mol Sci (2019) 20(8). Epub 20190424. doi: 10.3390/ijms20082009.

39. Larionova I, Tuguzbaeva G, Ponomaryova A, Stakheyeva M, Cherdyntseva N, Pavlov V, et al. Tumor-Associated Macrophages in Human Breast, Colorectal, Lung, Ovarian and Prostate Cancers. Front Oncol (2020) 10:566511. Epub 20201022. doi: 10.3389/fonc.2020.566511.

40. Luan Y, Mosheir E, Menon MC, Wilson D, Woytovich C, Ochando J, et al. Monocytic Myeloid-Derived Suppressor Cells Accumulate in Renal Transplant Patients and Mediate Cd4(+) Foxp3(+) Treg Expansion. Am J Transplant (2013) 13(12):3123–31. Epub 20130918. doi: 10.1111/ajt.12461.

41. Utrero-Rico A, Laguna-Goya R, Cano-Romero F, Chivite-Lacaba M, Gonzalez-Cuadrado C, Rodriguez-Sanchez E, et al. Early Posttransplant Mobilization of Monocytic Myeloid-Derived Suppressor Cell Correlates with Increase in Soluble Immunosuppressive Factors and Predicts Cancer in Kidney Recipients. Transplantation (2020) 104(12):2599–608. doi: 10.1097/TP.0000000000003179.

42. Bouwes Bavinck JN, Feltkamp MCW, Green AC, Fiocco M, Euvrard S, Harwood CA, et al. Human Papillomavirus and Posttransplantation Cutaneous Squamous Cell Carcinoma: A Multicenter, Prospective Cohort Study. Am J Transplant (2018) 18(5):1220–30. Epub 20171122. doi: 10.1111/ajt.14537.

43. Smola S, Trimble C, Stern PL. Human Papillomavirus-Driven Immune Deviation: Challenge and Novel Opportunity for Immunotherapy. Ther Adv Vaccines (2017) 5(3):69–82. Epub 20170705. doi: 10.1177/2051013617717914.

44. Lelios I, Stifter SA, Cecconi V, Petrova E, Lutz M, Cansever D, et al. Monocytes Promote UvLInduced Epidermal Carcinogenesis. European Journal of Immunology (2021) 51(7):1799–808. doi: 10.1002/eji.202048841.

45. Xiu B, Lin Y, Grote DM, Ziesmer SC, Gustafson MP, Maas ML, et al. Il-10 Induces the Development of Immunosuppressive Cd14(+)Hla-Dr(Low/-) Monocytes in B-Cell Non-Hodgkin Lymphoma. Blood Cancer J (2015) 5:e328. Epub 20150731. doi: 10.1038/bcj.2015.56.

