## Supporting Information for "Dampened inflammatory signalling and myeloid-derived suppressor-like cell accumulation reduces circulating monocytic HLA-DR density and associates with malignancy risk in long-term renal transplant recipients"

Supplementary Material

### 1 Supplementary Figures and Tables

#### 1.1 Supplementary Figures

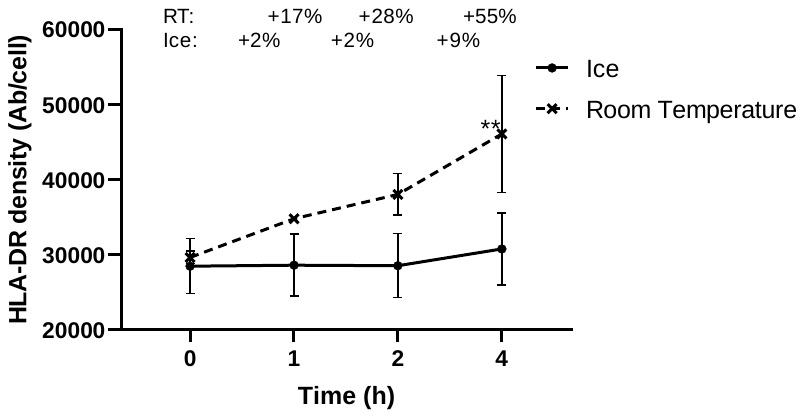

**Supplementary Figure 1: Effect of time and storage condition upon mHLA-DRd.** Blood was taken from healthy controls (n=3) and HLA-DR staining commenced after no delay (0) or 1-4 hours delay, with blood stored in the interim at room temperature (dashed line, crossed points) or on ice (solid line, circle points). Percentage values indicate the mean increase in mHLA-DRd from baseline with each condition. Timepoints were compared against mHLA-DRd at timepoint 0h by ANOVA followed by Dunnett’s multiple comparison test. ** adjusted p<0.01.

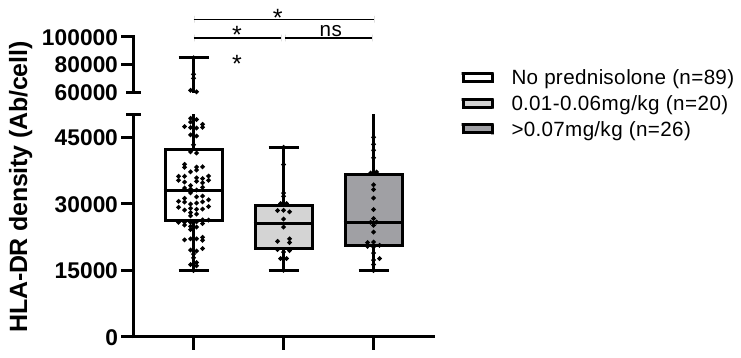

**Supplementary Figure 2: mHLA-DRd by prednisolone dose.** RTR were subdivided by weight-adjusted prednisolone dose. Groups were compared by one-way analysis of variance (ANOVA) followed by post-hoc Tukey’s multiple comparison testing. Box-whisker plot shows maximum and minimum values and quartile values for each group. * adjusted p <0.05, ** adjusted p <0.01, ‘ns’ not significant.

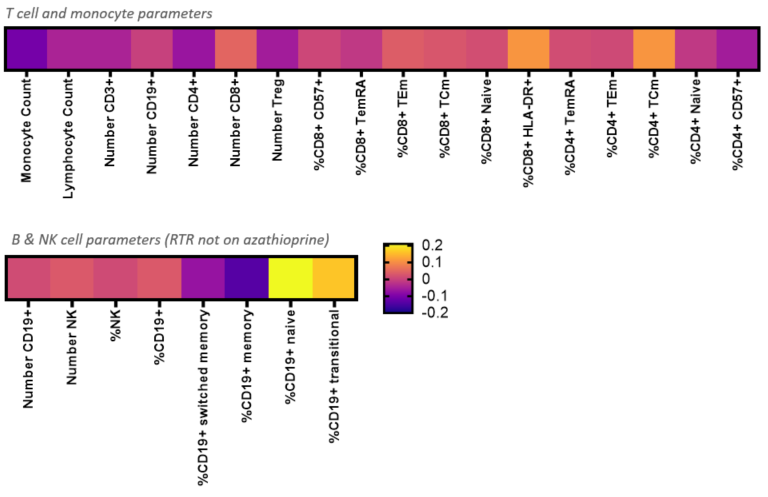

**Supplementary Figure 3: mHLA-DRd exhibits little correlation to other circulating immunological populations**. mHLA-DRd was correlated to other lymphocyte populations by Pearson’s correlation, with r values reported. All were non-significant.

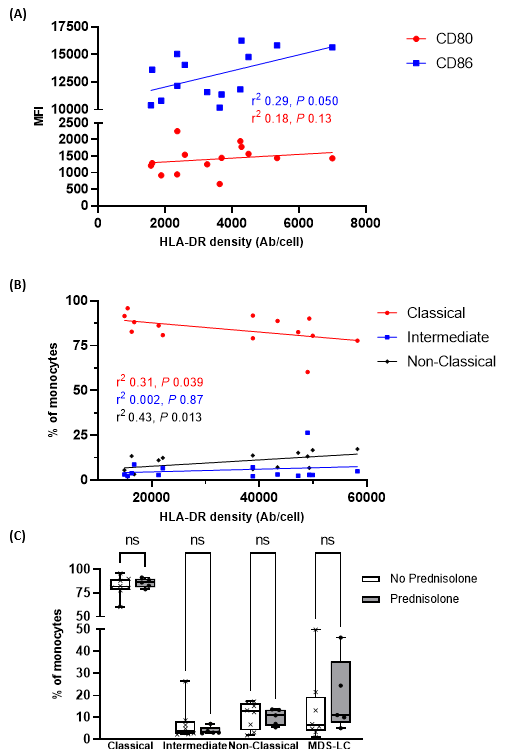

**Supplemental Figure 4: Correlation of mHLA-DRd and steroid use with monocyte phenotype.** (A) Correlation of mHLA-DRd with co-stimulatory receptor expression; (B) Correlation of major monocyte subpopulations with mHLA-DRd; (C) major monocyte populations stratified by corticosteroid (prednisolone) use at time of sampling. Correlations and significance are assessed using Spearman’s test (n=14) for (A) and (B), whilst (C) used Mann-Witney testing of each monocyte population.

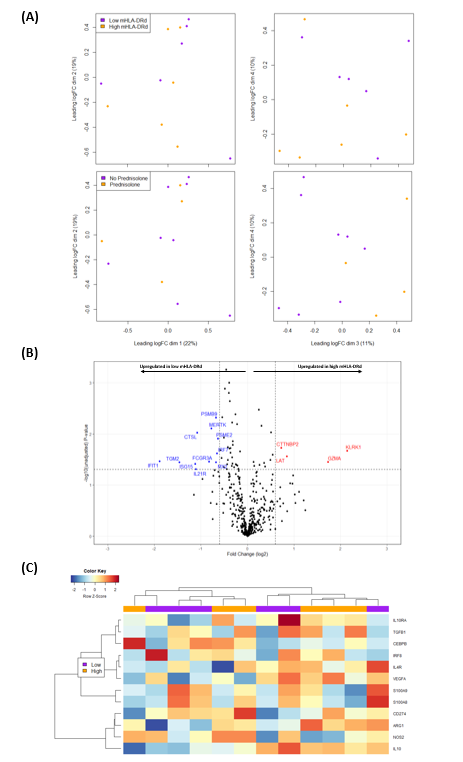

**Supplementary Figure 5: Differential gene expression.** (A) Principle component analysis stratified by mHLA-DRd expression (top panels) and prednisolone use at sampling (bottom panels). The first four dimensions are shown as multidimensional scaling plots. (B) Volcano plot illustrating differential gene expression in high and low mHLA-DRd groups. Genes demonstrating significant differential expression prior to multiple testing correction are highlighted (no genes demonstrated significance after correction). Note, y-axis indicates unadjusted *P* value. Dashed lines indicate log fold change of ±0.6 (x-axis) and unadjusted *P* value of 0.05 (y-axis). (C) Heatmap of selected genes previously associated with MDSC. Hierarchical clustered was unsupervised. Columns are labelled according to low or high mHLA-DRd.

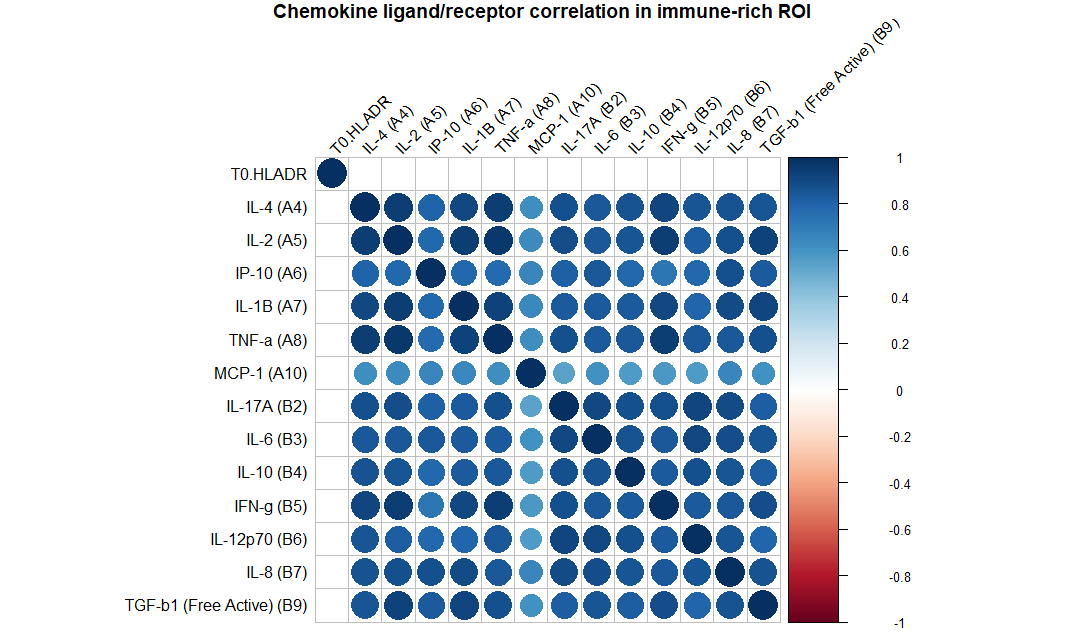

**Supplementary Figure 6: mHLA-DRd demonstrates no correlation to serum concentrations of a panel of cytokines**. Correlations were assessed using Spearman’s test and only those remaining significant after adjustment for multiple testing are shown.

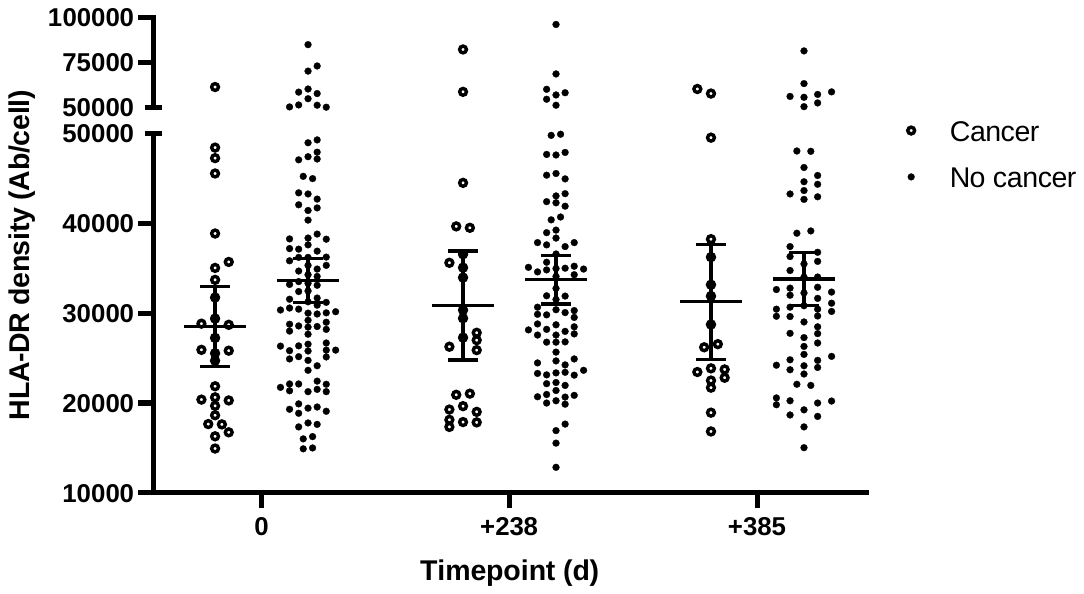

**Supplementary Figure 7: mHLA-DRd on serial sampling in those with and without malignancy development during follow-up.** Serial mHLA-DRd at three timepoints during study follow-up. Individual datapoints and mean and 95% confidence interval in those with and without malignancy development during follow-up are shown.

#### 1.2 Supplementary Tables

| **Supplier** | **Specificity and fluorochrome** | **Clone** |
| --- | --- | --- |
| eBioscience (now ThermoFisher Scientific), Paisley, UK | CD3-eFluor 450 | UCHT1 |
|  | Gamma-delta TCR – FITC | B1.1 |
|  | CD28-APC-eFluor780 | CD28.2 |
|  | CD57-PE | NK1.1 |
|  | CD38-APC | HIT2 |
|  | CD33-eFluor450 | P67.6 |
|  | CD20-APC-eFluor780 | 2H7 |
|  | 7-AAD | N/A |
| Beckman Coulter, Wycombe, UK | CD4-ECD | SFCI2T4D11 |
|  | HLA-DR-Krome Orange | Immu-357 |
|  | CD19-Krome Orange | J3-119 |
| BD Biosciences | CD8-PerCP-Cy5.5 | SK1 |
|  | CD16-PE-Cy7 | 3G8 |
|  | CD56-APC | B159 |
|  | CD27-FITC | M-T271 |
|  | CD127-PE | HIL-7R-M21 |
|  | CD25-PE-Cy7 | M-A251 |
|  | CD69-Alexa Fluor 700 | FN50 |
|  | CCR7-PE-Cy7 | 3D12 |
|  | CD24-PE | ML5 |
|  | Granzyme B-Alexa Fluor 700 | GB11 |
|  | IgD-PE-Cy7 | IA6-2 |
|  | CD86-PE | 2331 |
| Invitrogen (now ThermoFisher Scientific), Paisley, UK | CD45RA-APC | MEM-56 |
|  | CD80-FITC | 2D10.4 |
|  | CD14-APC-eFluor 780 | 61D3 |
| Biolegend, London, UK | FoxP3-Alexa Fluor 647 | 259D |
|  | CD19-Alexa Fluor 700 | HIB19 |
|  | CD56-Alexa Fluor 700 | 5.1H11 |
|  | CD3-Alexa Fluor 700 | UCHT1 |
|  | CD11b-APC | ICRF44 |

Antibodies (and clones) used were: D45RA-APC (MEM-56) from Invitrogen (Life Technologies, Paisley, UK); FoxP3-Alexa Fluor 647 (259D) from Biolegend, London, UK.

| **Factor** | **Beta, t(126)** | **p** |
| --- | --- | --- |
| Induction Therapy | -0.02, -0.22 | 0.83 |
| Age at recruitment | 0.03, 0.32 | 0.75 |
| On sirolimus at recruitment | 0.16, 1.89 | 0.06 |
| On prednisolone at recruitment | -0.30, -2.86 | 0.005 |
| Number of immunosuppressive agents at recruitment | -0.05, -0.49 | 0.62 |

**Supplementary Table 2: Multivariable linear regression assessing impact of clinical variables upon mHLA-DRd.** Regression was undertaken using simultaneous entry method using age as a pre-specified variable and other variables as identified upon univariable analysis.

|  | **Low mHLA-DRd (n=6)** | **High mHLA-DRd (n=6)** |
| --- | --- | --- |
| Age (yrs) | 66 (61 – 74) | 68 (54 – 75) |
| Male | 3 (50%) | 4 (66%) |
| Cumulative Immunosuppression duration (months) | 260 (103 – 364) | 287 (159 – 343) |
| mHLA-DRd (antibodies/cell) | 16521 (15398 – 21478) | 48084 (42225 – 51972) |
| Immunosuppression:  Prednisolone  Sirolimus  Calcineurin inhibitor  Azathioprine  Mycophenolate | 2 (33%)  0  5 (83%)  4 (66%)  1 (17%) | 2 (33%)  0  6 (100%)  5 (83%)  1 (17%) |
| Induction therapy:  None  Basiliximab  Alemtuzumab  Thymoglobulin | 3 (50%)  3 (50%)  0  0 | 5 (83%)  1 (17%)  0  0 |

**Supplementary Table 3: Details of samples used for monocyte enrichment and gene expression analysis.** Continuous variables are reported as median (interquartile range) whilst categorical variables are reported as n (percent).

| **ONTOLOGY** | **GO ID** | **Description** | **Set Size** | **Enrichment Score** | **NES** | **Raw *P* value** | **Adjusted *P* value** | **Q value** | **Rank** | **Leading Edge** | | | **Core enrichment** |
| --- | --- | --- | --- | --- | --- | --- | --- | --- | --- | --- | --- | --- | --- |
|  |  |  |  |  |  |  |  |  |  | **Tags** | **List** | **Signal** |  |
| MF | 0030545 | Receptor regulator activity | 112 | 0.688788490553563 | 1.85956190432751 | 5.94649900584774e-07 | 0.000273538954268996 | 0.000259142167202207 | 86 | 21% | 12% | 21% | IL1A/IL1B/CXCL2/CXCL3/CCL20/CXCL5/EGF/TNFSF4/CXCL1/IL10/CCL4/HBEGF/CXCL8/IL1RN/IL6/C5/CCL5/CCL3L1/PF4/VEGFA/OSM/BMP6/CCL3 |
| MF | 0030546 | Signaling receptor activator activity | 112 | 0.688788490553563 | 1.85956190432751 | 5.94649900584774e-07 | 0.000273538954268996 | 0.000259142167202207 | 86 | 21% | 12% | 21% | IL1A/IL1B/CXCL2/CXCL3/CCL20/CXCL5/EGF/TNFSF4/CXCL1/IL10/CCL4/HBEGF/CXCL8/IL1RN/IL6/C5/CCL5/CCL3L1/PF4/VEGFA/OSM/BMP6/CCL3 |
| MF | 0048018 | Receptor ligand activity | 112 | 0.688788490553563 | 1.85956190432751 | 5.94649900584774e-07 | 0.000273538954268996 | 0.000259142167202207 | 86 | 21% | 12% | 21% | IL1A/IL1B/CXCL2/CXCL3/CCL20/CXCL5/EGF/TNFSF4/CXCL1/IL10/CCL4/HBEGF/CXCL8/IL1RN/IL6/C5/CCL5/CCL3L1/PF4/VEGFA/OSM/BMP6/CCL3 |
| MF | 0005125 | Cytokine activity | 89 | 0.71048476824871 | 1.87049191665443 | 2.65919462962663e-06 | 0.000917422147221187 | 0.000869136771051651 | 86 | 24% | 12% | 24% | IL1A/IL1B/CXCL2/CXCL3/CCL20/CXCL5/TNFSF4/CXCL1/IL10/CCL4/CXCL8/IL1RN/IL6/C5/CCL5/CCL3L1/PF4/VEGFA/OSM/BMP6/CCL3 |
| MF | 0048020 | CCR chemokine receptor binding | 28 | 0.880768387583542 | 1.88081378242469 | 1.00469566581571e-05 | 0.00277296003765137 | 0.00262701477251182 | 86 | 21% | 12% | 20% | CCL20/CCL4/CCR2/CCL5/CCL3L1/CCL3 |
| BP | 0040011 | Locomotion | 273 | 0.507745168556908 | 1.50387178547316 | 1.67263570041186e-05 | 0.00384706211094729 | 0.00364458515773953 | 105 | 20% | 14% | 27% | KLRK1/IL1B/PTGS2/NFATC2/ANGPT1/CXCL2/GPR183/CXCL3/PROK2/CCL20/CXCL5/CXCR4/ETS1/ADORA2A/EPHB6/CYP1B1/EGF/PLAUR/CXCL1/IL10/FGFR1/NLRP3/CCL4/THBD/HBEGF/CXCL8/S100A9/TNFAIP3/SELP/S100A8/CCR2/OLR1/IL6/C5/NR4A2/CCL5/CCR4/ALCAM/CCL3L1/SIRPA/PF4/VEGFA/CLIC4/CCL3/CD99/ICAM1/PRKCI/CSF3R/ID1/PROS1/IL1R1/FLT1/MAPK14/ADAM8 |
| MF | 0098772 | Molecular function regulator | 195 | 0.554444033944999 | 1.58292960243998 | 2.67458023974775e-05 | 0.005272743901217 | 0.00499523106431084 | 120 | 19% | 16% | 22% | IL1A/IL1B/RGS1/CXCL2/CXCL3/SOCS3/CCL20/CXCL5/EGF/TNFSF4/CXCL1/IL10/CCL4/HBEGF/CXCL8/ID2/ALOX5AP/IL1RN/SERPINB2/IL6/C5/CCL5/CCL3L1/SIRPA/PF4/VEGFA/OSM/BMP6/CCL3/ID1/PROS1/BIRC3/BTG2/PRDX3/HMGB1/TGFA/RAF1 |
| BP | 0016477 | Cell migration | 248 | 0.514935764684132 | 1.51267632924005 | 6.68086472862491e-05 | 0.0104142118044655 | 0.0098660953937042 | 100 | 19% | 14% | 25% | KLRK1/IL1B/PTGS2/NFATC2/ANGPT1/CXCL2/GPR183/CXCL3/CCL20/CXCL5/CXCR4/ETS1/CYP1B1/EGF/CXCL1/IL10/FGFR1/NLRP3/CCL4/THBD/HBEGF/CXCL8/S100A9/TNFAIP3/SELP/S100A8/CCR2/OLR1/IL6/C5/NR4A2/CCL5/CCR4/CCL3L1/SIRPA/PF4/VEGFA/CLIC4/CCL3/CD99/ICAM1/PRKCI/CSF3R/ID1/PROS1/IL1R1/FLT1 |
| BP | 0006928 | Movement of cell or subcellular component | 265 | 0.503337213201213 | 1.47976045777804 | 6.79187726378188e-05 | 0.0104142118044655 | 0.0098660953937042 | 100 | 18% | 14% | 25% | KLRK1/IL1B/PTGS2/NFATC2/ANGPT1/CXCL2/GPR183/CXCL3/CCL20/CXCL5/CXCR4/ETS1/EPHB6/CYP1B1/EGF/CXCL1/IL10/FGFR1/NLRP3/CCL4/THBD/HBEGF/CXCL8/S100A9/TNFAIP3/SELP/S100A8/CCR2/OLR1/IL6/C5/NR4A2/CCL5/CCR4/ALCAM/CCL3L1/SIRPA/PF4/VEGFA/CLIC4/CCL3/CD99/ICAM1/PRKCI/CSF3R/ID1/PROS1/IL1R1/FLT1 |
| MF | 0005126 | Cytokine receptor binding | 106 | 0.634329903896816 | 1.69380879852863 | 7.78163216400592e-05 | 0.0107386523863282 | 0.0101734601554688 | 86 | 20% | 12% | 20% | IL1A/IL1B/CXCL2/CXCL3/CCL20/CXCL5/TNFSF4/CXCL1/IL10/CCL4/CXCL8/IL1RN/CCR2/IL6/C5/CCL5/CCL3L1/PF4/VEGFA/OSM/CCL3 |
| MF | 0042379 | Chemokine receptor binding | 43 | 0.768586877819508 | 1.77660546099612 | 9.3092482340253e-05 | 0.0116788750572317 | 0.0110641974226406 | 86 | 30% | 12% | 28% | CXCL2/CXCL3/CCL20/CXCL5/CXCL1/CCL4/CXCL8/CCR2/C5/CCL5/CCL3L1/PF4/CCL3 |
| BP | 0048870 | Cell motility | 252 | 0.506152064012107 | 1.48629434665459 | 0.000128656237129335 | 0.0136573544029602 | 0.0129385462764886 | 100 | 19% | 14% | 25% | KLRK1/IL1B/PTGS2/NFATC2/ANGPT1/CXCL2/GPR183/CXCL3/CCL20/CXCL5/CXCR4/ETS1/CYP1B1/EGF/CXCL1/IL10/FGFR1/NLRP3/CCL4/THBD/HBEGF/CXCL8/S100A9/TNFAIP3/SELP/S100A8/CCR2/OLR1/IL6/C5/NR4A2/CCL5/CCR4/CCL3L1/SIRPA/PF4/VEGFA/CLIC4/CCL3/CD99/ICAM1/PRKCI/CSF3R/ID1/PROS1/IL1R1/FLT1 |
| BP | 0051674 | Localization of cell | 252 | 0.506152064012107 | 1.48629434665459 | 0.000128656237129335 | 0.0136573544029602 | 0.0129385462764886 | 100 | 19% | 14% | 25% | KLRK1/IL1B/PTGS2/NFATC2/ANGPT1/CXCL2/GPR183/CXCL3/CCL20/CXCL5/CXCR4/ETS1/CYP1B1/EGF/CXCL1/IL10/FGFR1/NLRP3/CCL4/THBD/HBEGF/CXCL8/S100A9/TNFAIP3/SELP/S100A8/CCR2/OLR1/IL6/C5/NR4A2/CCL5/CCR4/CCL3L1/SIRPA/PF4/VEGFA/CLIC4/CCL3/CD99/ICAM1/PRKCI/CSF3R/ID1/PROS1/IL1R1/FLT1 |
| MF | 0001664 | G protein-coupled receptor binding | 57 | 0.711553908355034 | 1.74416976741888 | 0.000201336731165541 | 0.0198460492148891 | 0.0188015203088423 | 88 | 28% | 12% | 27% | CXCL2/CXCL3/PROK2/CCL20/CXCL5/ADORA2A/CXCL1/CCL4/CXCL8/CCR2/C5/CCL5/CCL3L1/PF4/CCL3/FCN1 |
| MF | 0008009 | Chemokine activity | 41 | 0.762236891776592 | 1.73564199384638 | 0.000243965832996036 | 0.0224448566356353 | 0.0212635483916545 | 86 | 29% | 12% | 27% | CXCL2/CXCL3/CCL20/CXCL5/CXCL1/CCL4/CXCL8/C5/CCL5/CCL3L1/PF4/CCL3 |
| CC | 0005788 | Endoplasmic reticulum lumen | 31 | 0.804895769043644 | 1.75948690376638 | 0.000346977266711301 | 0.0270569599522001 | 0.0256329094284001 | 60 | 10% | 8% | 9% | PTGS2/PLAUR/IL6 |
| BP | 0065009 | Regulation of molecular function | 301 | 0.461145860529582 | 1.37169011661993 | 0.000376820199170817 | 0.0270569599522001 | 0.0256329094284001 | 86 | 15% | 12% | 23% | KLRK1/GZMA/IL1A/IL1B/RGS1/ANGPT1/LAT/SOCS3/PROK2/CCL20/CXCR4/DUSP2/ADORA2A/EPHB6/CYP1B1/EGF/PLAUR/TNFSF4/CD36/CXCL1/IL10/FGFR1/NLRP3/CCL4/PTX3/HBEGF/FCGR2B/MYC/S100A9/TNFAIP3/ID2/ALOX5AP/S100A8/CCR2/SERPINB2/IL6/C5/NR4A2/CCL5/NFKBIA/CCL3L1/SIRPA/GADD45B/VEGFA/RIPK2/CCL3 |
| BP | 0006935 | Chemotaxis | 154 | 0.546588390966672 | 1.52357663493674 | 0.000389513364243902 | 0.0270569599522001 | 0.0256329094284001 | 105 | 21% | 14% | 23% | KLRK1/ANGPT1/CXCL2/GPR183/CXCL3/PROK2/CCL20/CXCL5/CXCR4/EPHB6/PLAUR/CXCL1/IL10/FGFR1/CCL4/HBEGF/CXCL8/S100A9/S100A8/CCR2/IL6/C5/CCL5/CCR4/ALCAM/CCL3L1/PF4/VEGFA/CCL3/CSF3R/FLT1/MAPK14/ADAM8 |
| BP | 0042330 | Taxis | 154 | 0.546588390966672 | 1.52357663493674 | 0.000389513364243902 | 0.0270569599522001 | 0.0256329094284001 | 105 | 21% | 14% | 23% | KLRK1/ANGPT1/CXCL2/GPR183/CXCL3/PROK2/CCL20/CXCL5/CXCR4/EPHB6/PLAUR/CXCL1/IL10/FGFR1/CCL4/HBEGF/CXCL8/S100A9/S100A8/CCR2/IL6/C5/CCL5/CCR4/ALCAM/CCL3L1/PF4/VEGFA/CCL3/CSF3R/FLT1/MAPK14/ADAM8 |
| BP | 006954 | Inflammatory response | 229 | 0.496776813747341 | 1.44348329820547 | 0.000392129854379712 | 0.0270569599522001 | 0.0256329094284001 | 114 | 24% | 16% | 30% | IL1A/IL1B/PTGS2/CXCL2/LAT/CXCL3/SOCS3/PROK2/MAPK13/CCL20/CXCL5/CXCR4/ETS1/ADORA2A/TNFSF4/CD36/CXCL1/IL10/NLRP3/CCL4/PTX3/FCGR2B/CXCL8/S100A9/TNFAIP3/IL1RN/SELP/S100A8/CCR2/OLR1/IL6/C5/CCL5/CCR4/NFKBIA/CCL3L1/PTGS1/SIRPA/PF4/OSM/BMP6/RIPK2/CCL3/PTGER2/ICAM1/IL1R1/CD180/TLR10/TLR5/MAPK14/ADAM8/BIRC3/AOAH/LIPA/CXCR2 |

**Supplementary Table 4: Results of enrichment analysis using Gene Ontology knowledgebase.** MF, molecular function; BP, biological process; CC, cellular component. Result are listed in order of ascending adjusted *P* value.
